# Inborn errors of immunity: manifestation, treatment, and outcome – an ESID registry 1994-2024 report on 30,628 patients

**DOI:** 10.1101/2025.02.20.25322586

**Authors:** Gerhard Kindle, Mickaël Alligon, Michael H. Albert, Matthew Buckland, J. David Edgar, Benjamin Gathmann, Sujal Ghosh, Antonios Gkantaras, Alexandra Nieters, Claudio Pignata, Peter Robinson, Stephan Rusch, Catharina Schuetz, Svetlana Sharapova, Benjamin Shillitoe, Fabio Candotti, Andrew J. Cant, Jean-Laurent Casanova, Amos Etzioni, Alain Fischer, Isabelle Meyts, Luigi D. Notarangelo, Martine Pergent, C. I. Edvard Smith, ESID Registry Working Party, Lennart Hammarström, Bodo Grimbacher, Mikko Seppänen, Nizar Mahlaoui, Stephan Ehl, Markus G. Seidel

**Affiliations:** Institute for Immunodeficiency, Center for Chronic Immunodeficiency, Medical Center, University of Freiburg, Faculty of Medicine, University of Freiburg, Freiburg, Germany; Centre for Biobanking FREEZE, Medical Center-University of Freiburg, Faculty of Medicine, University of Freiburg, Freiburg, Germany; French National Reference Center for Primary Immune Deficiencies (CEREDIH), Necker Enfants Malades University Hospital, Assistance Publique-Hôpitaux de Paris (AP-HP), Paris, France; Department of Pediatrics, Dr. von Hauner Children’s Hospital, Ludwig-Maximilians Universität München, Munich Germany; Barts Health National Health Service Trust, London, United Kingdom; Molecular and Cellular Immunology Section, Immunity and Inflammation Department, Great Ormond Street Institute of Child Health, London, United Kingdom; Department of Immunology, St James’s Hospital, & School of Medicine, Trinity College Dublin, Ireland; Department of Pediatric Oncology, Hematology and Clinical Immunology, Medical Faculty, Center of Child and Adolescent Health, Heinrich-Heine-University and University Hospital, Duesseldorf, Germany; Pediatric Immunology and Rheumatology Referral Center, 1st Department of Pediatrics, Aristotle University of Thessaloniki, Hippokration General Hospital, Thessaloniki, Greece; Department of Translational Medical Sciences, Federico II University, Naples, Italy; Berlin Institute of Health, Charité - Universitätsmedizin Berlin, Berlin, Germany; The Jackson Laboratory for Genomic Medicine, Farmington, CT, USA; Department of Pediatrics and University Center for Rare Diseases, Medizinische Fakultät Carl Gustav Carus, Technische Universität, Dresden, Germany; German Center for Child and Adolescent Health (DZKJ), partner site Leipzig/Dresden, Dresden, Germany; Research Department, Belarusian Research Center for Pediatric Oncology, Hematology and Immunology, Minsk Region, Belarus; Paediatric Immunology and Infectious Diseases, Sheffield Children’s NHS Foundation Trust, Sheffield, United Kingdom; Division of Immunology and Allergy, Lausanne University Hospital and University of Lausanne, Lausanne, Switzerland; Translational and Clinical Research Institute, Newcastle University; Department of Paediatric Immunology and Infectious Diseases, Great North Children’s Hospital, Newcastle upon Tyne, England, UK; Laboratory of Human Genetics of Infectious Diseases, INSERM U1163, Necker Hospital for Sick Children, Paris, France; Imagine Institute, University Paris Cité, Paris, France; St Giles Laboratory of Human Genetics of Infectious Diseases, Rockefeller Branch, The Rockefeller University, New York, NY, USA; Howard Hughes Medical Institute, New York, NY, USA; Department of Pediatrics, Necker Hospital for Sick Children, AP-HP, Paris, France; Faculty of Medicine, Technion, Haifa, Israel; Unité d’immunologie pédiatrique, Hôpital Necker Enfants Malades, Paris; Department of Pediatrics, Department of Microbiology, Immunology and Transplantation, University Hospitals Leuven, KU Leuven, Leuven, Belgium, ERN-RITA Core Center; Laboratory of Clinical Immunology and Microbiology, National Institute of Allergy and Infectious Diseases, National Institutes of Health, Bethesda, MD, USA; International Patient Organisation for Primary Immunodeficiencies (IPOPI), Brussels, Belgium; Department of Laboratory Medicine, Clinical Immunology, Karolinska Institutet, Stockholm; Department of Infectious Diseases, Karolinska University Hospital, Stockholm, Sweden; Karolinska ATMP Center, Karolinska Institutet, Karolinska University Hospital, Stockholm, Sweden; Division of Immunology, Dept of Medical Biochemistry and Biophysics, Karolinska Institutet, Stockholm, Sweden; Clinic of Rheumatology and Clinical Immunology, Center for Chronic Immunodeficiency (CCI), Medical Center, Faculty of Medicine, Albert-Ludwigs-University of Freiburg, Germany; DZIF - German Center for Infection Research, Satellite Center Freiburg, Germany; CIBSS - Centre for Integrative Biological Signalling Studies, Albert-Ludwigs University, Freiburg, Germany; RESIST - Cluster of Excellence 2155 to Hanover Medical School, Satellite Center Freiburg, Germany; Rare Disease Center and Pediatric Research Center, Children and Adolescents, University of Helsinki, HUS Helsinki University Hospital, ERN-RITA Core Center, RITAFIN, Helsinki; Translational Immunology Research Program, University of Helsinki, Finland; Pediatric Immuno-Hematology and Rheumatology Unit, Necker Enfants Malades University Hospital, Assistance Publique-Hôpitaux de Paris (AP-HP), Paris, France; Styrian Children’s Cancer Research Unit for Cancer and Inborn Errors of the Blood and Immunity in Children; Division of Pediatric Hematology and Oncology, Department of Pediatric and Adolescent Medicine, Medical University of Graz, Graz, Austria

**Keywords:** Inborn error of immunity (IEI), primary immunodeficiency (PID), primary immune disorder (PID), online database, patient registry, overall survival

## Abstract

The *European Society for Immunodeficiencies* patient registry (ESID-R), established in 1994, is one of the world’s largest databases on inborn errors of immunity (IEI). IEI are genetic disorders predisposing patients to infections, autoimmunity, inflammation, allergies and malignancies. Treatments include antimicrobial therapy, immunoglobulin replacement, immune modulation, stem cell transplantation and gene therapy. Data from 194 centers in 33 countries capture clinical manifestations and treatments from birth onward, with annually expected updates. This report reviews the ESID-R’s structure, data content, and impact.

The registry includes 30,628 patient datasets (aged 0–97.9 years; median follow-up: 7.2 years; total 825,568.2 patient-years), with 13,550 cases in 15 sub-studies. It has produced 84 peer-reviewed publications (mean citation rate: 95). Findings include real-world observations of IEI diagnoses, genetic causes, clinical manifestations, treatments, and survival trends. The ESID-R fosters global collaboration, advancing IEI research and patient care. This report highlights the key role of the multi-national ESID-R, led by an independent medical society, in evidence-based discovery.

**Summary:** Worldwide, most patient registries for inborn errors of immunity are national, with limited geographical and temporal scope. This 30-year ESID registry analysis of 30,628 patients’ longitudinal datasets enables robust epidemiological studies on natural disease courses including diagnosis, treatment, and survival, supporting expanded newborn screening and future AI applications in IEI research.

## Introduction

Inborn errors of immunity (IEI) are genetic disorders affecting immunity. They almost invariably increase the susceptibility to infections and may cause autoimmunity, inflammation, allergy, and predispose individuals to malignancy(1-6). IEI can manifest at any age and are characterized by a spectrum of symptoms related to impaired or uncontrolled immune responses. These often have a serious impact on the health and quality of life of those affected. Many IEI are rare diseases, including ultra-rare and hyper-rare forms (*i.e.*, frequently <<1/2000 persons affected). Thus, an international strategy was needed to collect a meaningful number of datapoints for cross-sectional analyses and longitudinal clinical observations of natural courses of specific IEI.

The *European Society for Immunodeficiencies* registry (ESID-R) was founded in 1994 by ESID, a non-profit medical specialist society, creating a central database for primary immunodeficiencies. These are currently referred to as IEI or primary immune disorders (PID), differentiating them from secondary immune disorders (SID)(7). The ESID-R mainly serves to contain, store, and enable the analysis of IEI/PID data to improve the understanding of these diseases and their underlying immunobiology. Historically, the ESID-R was operated for the first 10 years as a hard copy-based database, and data were submitted by fax to the first chairs of the ESID-R, Lennart Hammarström and Mohammad Abedi in Huddinge, Sweden. From 2000–2004, Bodo Grimbacher with coworkers in Freiburg, Germany, developed and implemented the first web-based version of the ESID-R(8 9). In 2014, another major revision was carried out, headed by Stephan Ehl, Freiburg. The two key drivers of this second re-design were the goal to allow registry participation at three different levels according to center resources and the need to improve data quality so that a high level of confidence in the accuracy of the data could be inferred for clinical application and publication. In addition to inbuilt quality checks, the migration of existing data to the third version of the ESID-R and the entry of new patient data since then required a manual validation process of all included patients lacking a genetic diagnosis to meet the simultaneously developed clinical working criteria for IEI/PID diagnoses, compiled and published by a group of experts(10). In late 2024, ESID decided to move the ESID-R technical and physical foundation from the Medical Center of the University of Freiburg to a commercial clinical trials operator (Castor-edc, NL) to improve data-, system-, and access security. Maintaining the three-level study structure and the mandatory diagnosis validation process, this currently new, fourth version of the ESID-R is expected to facilitate the generation of data modules by research groups, allowing decentralized sub-study (eCRF) programming, independent data exports for center or sub-study analyses, automated center dashboards and add-on features such as patient reporting.

Here, we present major findings from clinical observations of 30,628 patients, based on the current ESID-R dataset. These findings indicate epidemiologically relevant disease distributions and the calculated prevalence of diagnoses in registered patients, their clinical manifestations, diagnostic delays, treatment, disease course and survival probabilities across all IEI/PID categories. Furthermore, we describe the organizational and technical evolution of the ESID-R, its relationship with the international registry landscape, and its role as a research (sub-)study platform. These data are of the utmost relevance to anyone affected by immune disorders or involved in patient care, management and therapy, drug and policy development for patients with IEI/PID around the world.

## Methods

### Technical background and operating mode of the ESID-R

The study protocol, the patient informed consent template and the center data transfer agreement plus amendments thereof were approved by the IRB and the data protection officer at the Medical University of Freiburg, Germany (Albert-Ludwigs-University). A substantial amendment was approved by the Medical University of Graz, Austria (24-334 ex 11/12, IRB00002556) and implemented in 2024. Details of the operational structure and technical background are found in the *Supplementary Material*.

### Data and statistical analyses

The study population included all patients with IEI/PID recorded in the ESID-R on March 19, 2024. Patients considered as discharged (*n*=538), patients with secondary immunodeficiency who were documented as part of one national subregistry only (*n*=417), or without definitive IEI diagnosis (n=306) were excluded from the study, leaving 30,628 patients. IEI were classified using the latest classification of the *International Union of Immunological Societies* (IUIS)(6), with abbreviations of the category names using, *e.g.*, “PIRD”, typically referred to as primary immune regulatory diseases, as synonym for “Diseases of immune dysregulation” (category *IV*; *i.e.*, including familial hemophagocytic lymphohistiocytosis, FHL). The statistical analyses were conducted using R (version 4.3.2, Vienna, Austria). Continuous and categorical variables were reported using the medians and interquartile ranges, raw effectives, and percentages, respectively. Survival probabilities were estimated by applying the cumulative incidence function. Overall survival (OS) was defined as the time between birth and death from any cause. Curative therapies, such as allogeneic hematopoietic stem cell transplantation and gene therapy, were considered as competing events. Survival was analyzed using the R package *survival* version 3.8-3 as described previously(11). As only patients with an eligible diagnosis are included, they are not considered to be at risk of dying before they are diagnosed(11). The prevalence was calculated based on the European population in 2019 (R package *rnaturalearth* version 1.0.1).

## Results

On the end date chosen for inclusion, the ESID-R contained data for 30,628 IEI/PID patients from 194 participating centers in 33 European and other, mostly neighboring countries. There was a steady increase in the registration of patients over time (supplementary figure 1) apart from a temporary decline in patient numbers and, to a lesser extent, in center numbers shortly after 2014, due to structural platform changes in 2014. These required centers to verify their patients to improve the accuracy of existing data. This allowed us to derive patient distribution by country and the minimal prevalence of PID/IEI (supplementary table 1 and supplementary figure 2).

### Patient characteristics

Figure 1 shows the ages at onset, clinical and genetic diagnoses and main clinical manifestations that led to diagnosis. A steady decline in new diagnoses was observed with age, although a small second peak and plateau were seen in adulthood, and a trend for earlier identification of genetic diagnoses was observed. The delay from the onset of symptoms to clinical diagnosis and between clinical and genetic diagnosis is shown by year of clinical or genetic diagnosis, *i.e.*, the endpoint of each delay, over the last 20 years in supplementary figure 1C. As reported previously(12), the majority of 80.3% of patients have infections on their way to diagnosis, but only in 61.8% as sole recorded manifestation of IEI, followed by features of immune dysregulation in 11.1% and syndromic manifestations in 7.3% as sole clinical presentation at diagnosis (*n*=15,360; 2,746; 1,803, respectively; figure 1B). Malignancies were among the first manifestations of IEI in 479 patients, and the only initial manifestation in 117 patients (0.5%; figure 1B).

**Figure 1.**
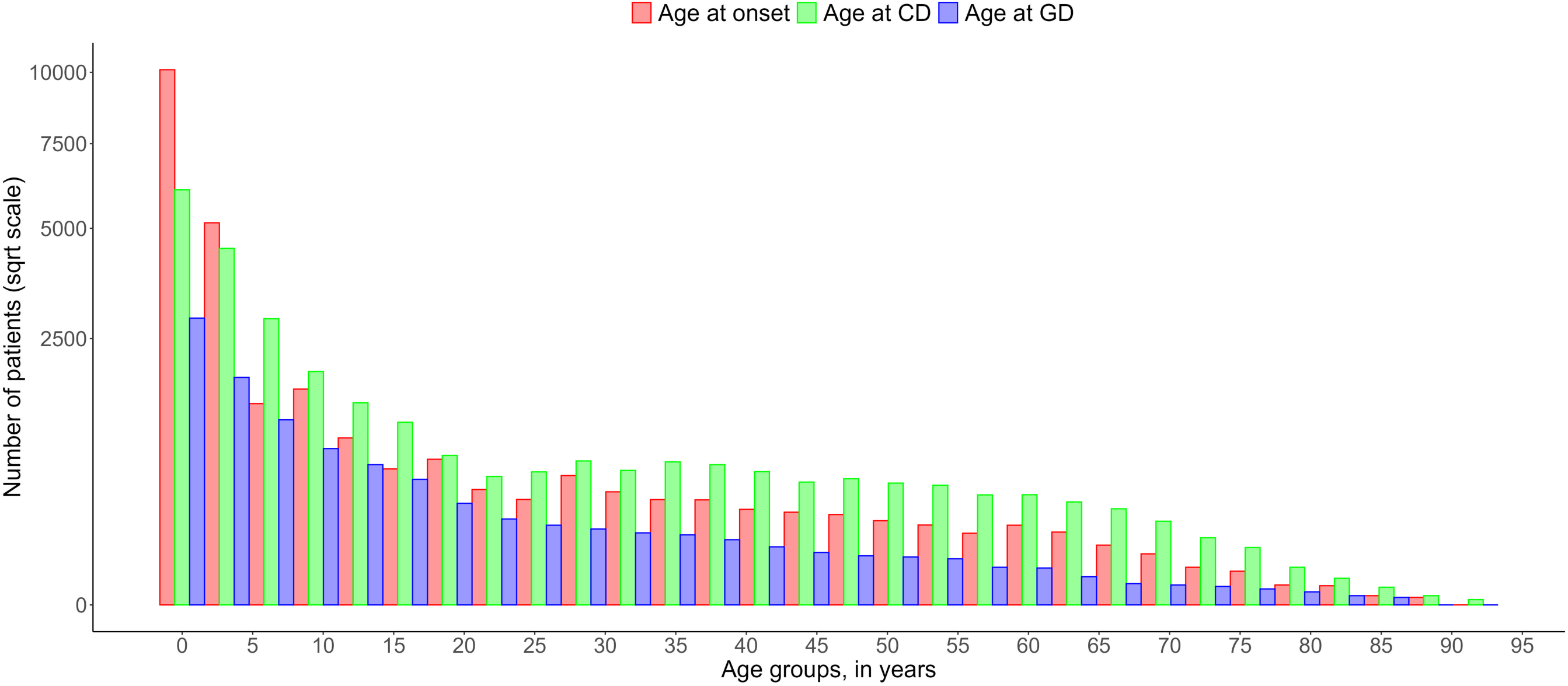
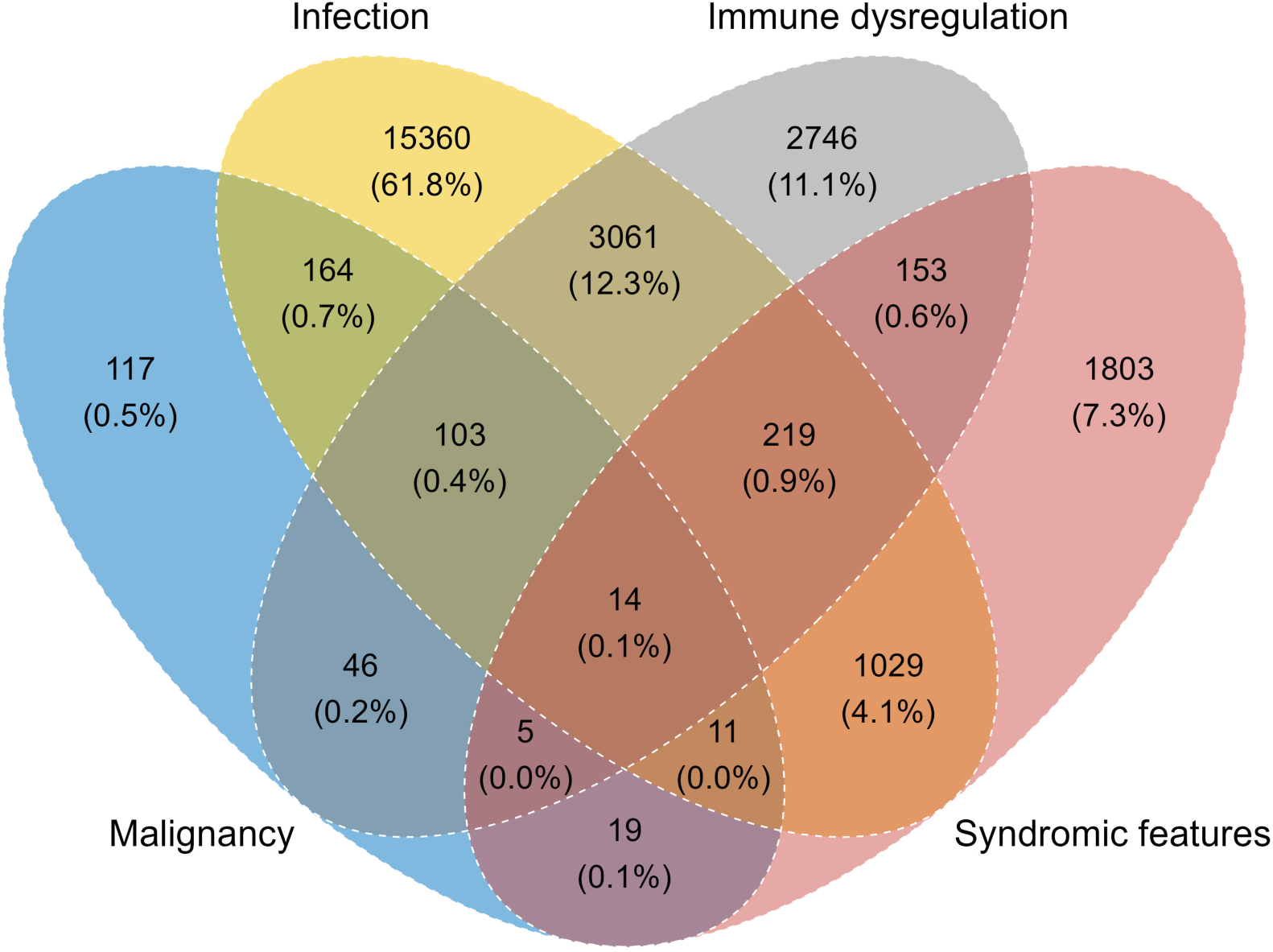
Ages and main manifestations at onset of inborn errors of immunity (IEI) or primary immune disorders (PID). ***A,*** age groups of age at disease onset, clinical diagnosis of IEI/PID, and of genetic diagnosis are shown. ***B,*** Venn diagram of main manifestations of IEI/PID with absolute patient numbers and proportions.

Table 1 shows the main patient demographic and diagnosis categories, comorbidities such as malignancy, COVID-19 and living status. The interactive supplementary figure 3 shows the patient numbers and their distribution according to their IUIS classification, disease name, and genetic diagnosis (https://esid.org/html-pages/Suppl%20Fig%203_ESID_30k_sunburst_PID.html). About half of all patients suffer from primary antibody deficiencies (PAD; *n*=15,123), followed in descending order, by combined immunodeficiencies with syndromic features (“syndromic”; *n*=4,239), phagocytic disorders (“phagocyte”; *n*=2,548), combined immunodeficiencies (CID; *n*=2,531), and primary diseases of immune dysregulation (PIRD; *n*=2,171; table 1 and supplementary figure 3). Table 1 presents the ages at onset, clinical diagnosis, genetic confirmation, and last follow-up, and the diagnostic delay. The proportion of patients reported to have suffered from malignancies was 8.9% (*n*=1,783). Malignancies were reported as occurring in all subgroups of patients with IEI/PID with moderately varying frequencies, corroborating the notion that IEI/PID are cancer predisposition syndromes. The proportion of patients reported to have been affected by COVID-19 was highest in PAD (35.9%; total cohort: 29.7%). More than half of CID patients received curative therapy (52.6%), followed by those with bone marrow failure syndromes (BMF; 39.8%), PIRD, and phagocyte disorders (30.3 and 25.8%, respectively). Supplementary Table 2 shows the causes of death.

**Table 1.**
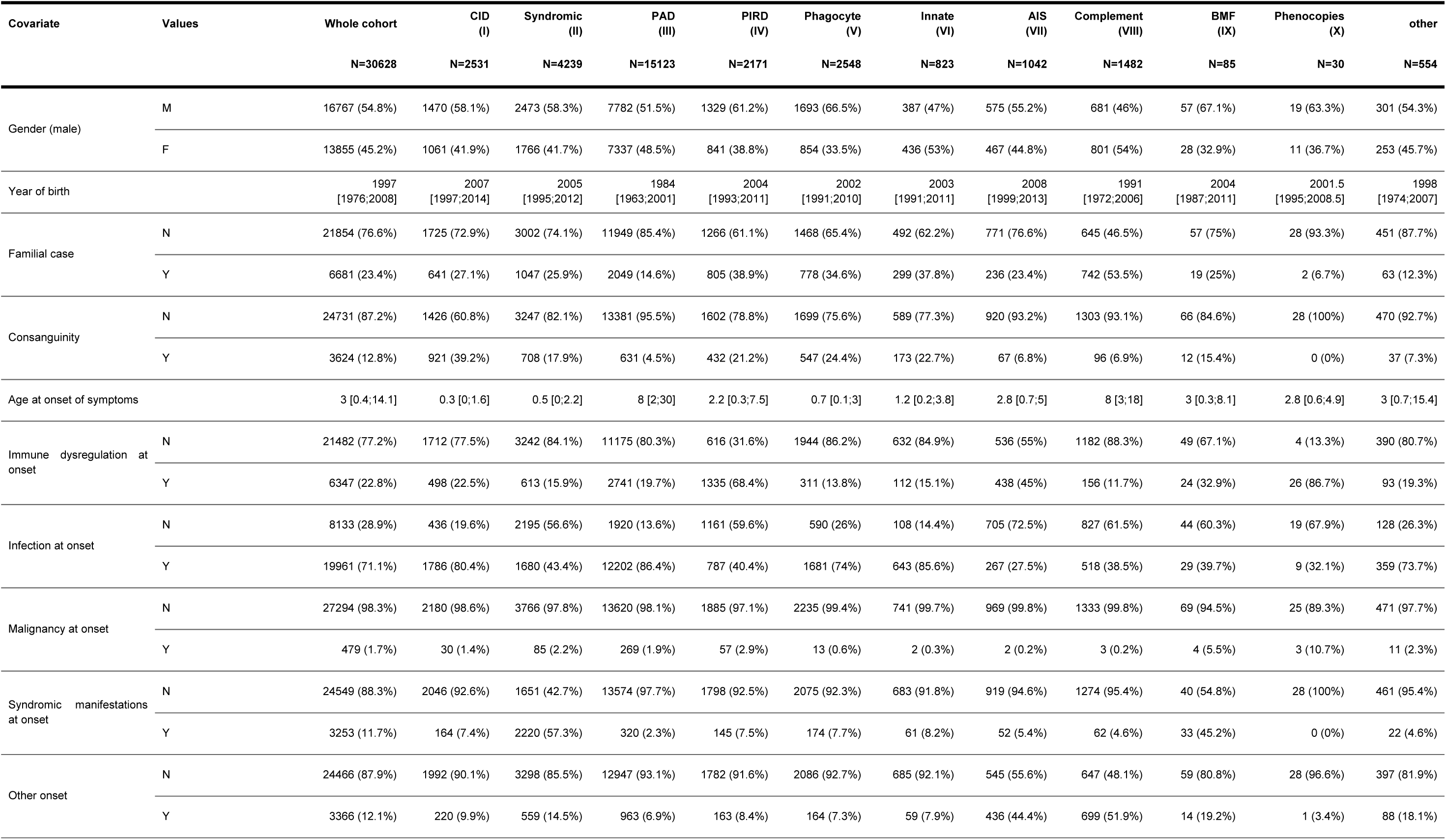
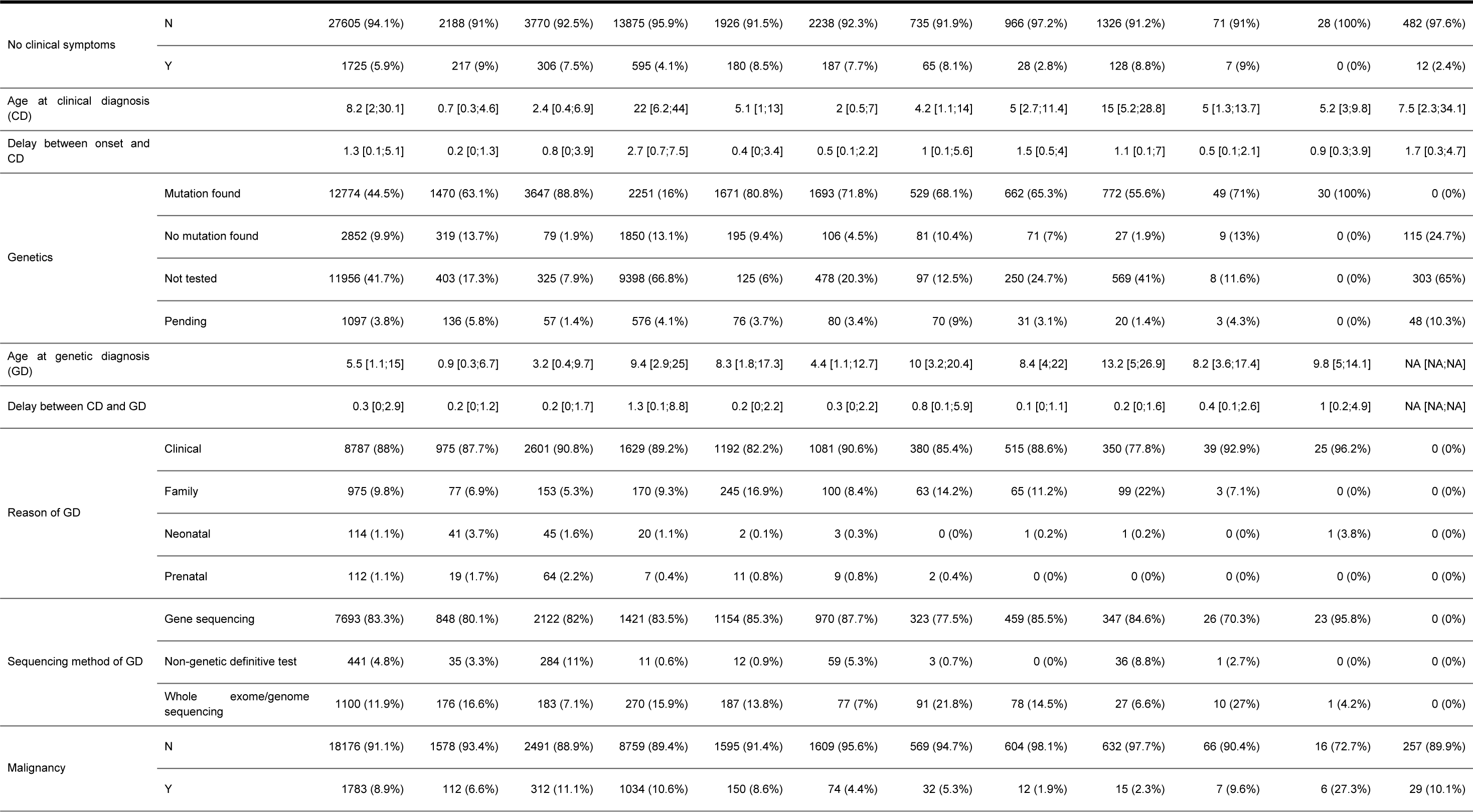
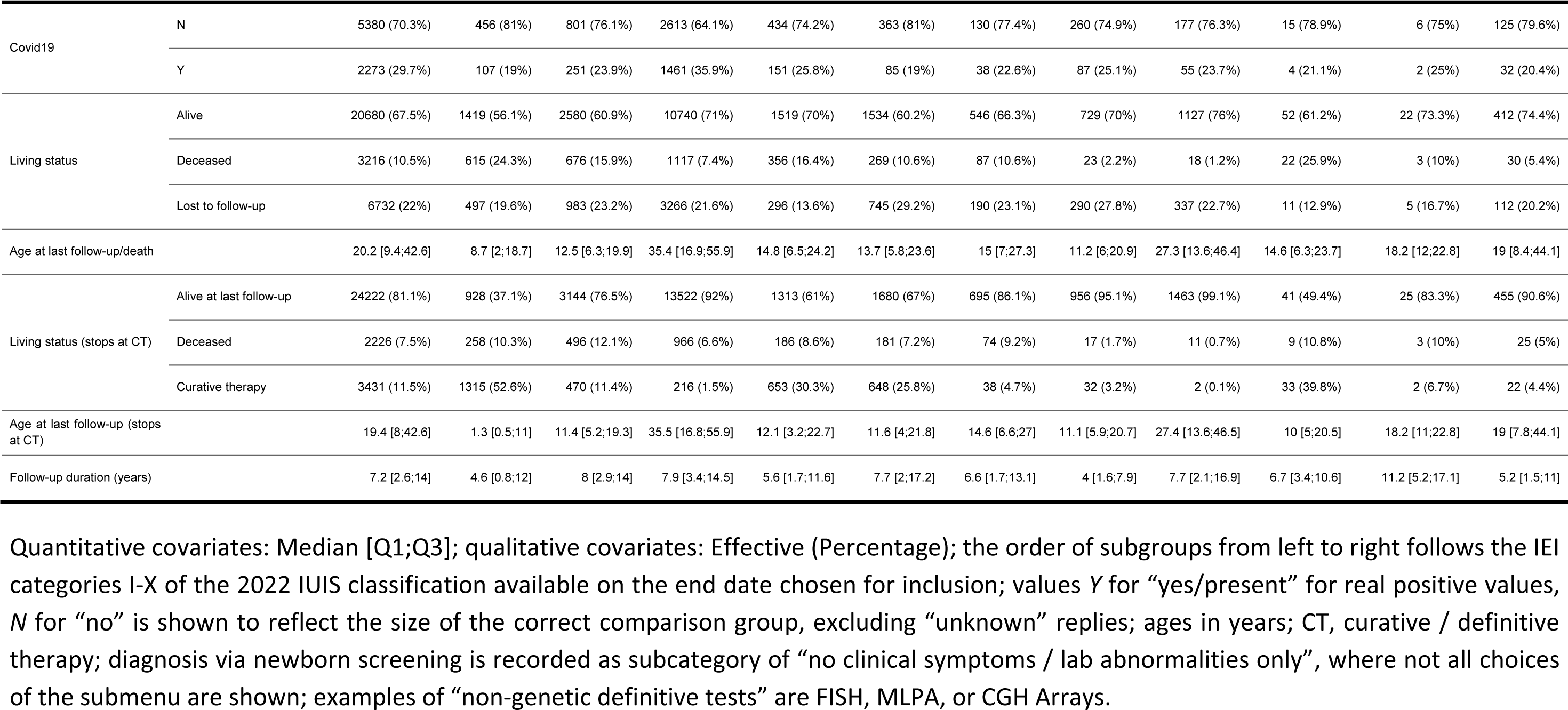
*European Society for Immunodeficiencies* (ESID) registry patient characteristics.

| Covariate | Values | Whole cohort | CID<br>(I) | Syndromic<br>(II) | PAD<br>(III) | PIRD<br>(IV) | Phagocyte<br>(V) | Innate<br>(VI) | AIS<br>(VII) | Complement<br>(VIII) | BMF<br>(IX) | Phenocopies<br>(X) | other |
| --- | --- | --- | --- | --- | --- | --- | --- | --- | --- | --- | --- | --- | --- |
|  |  | N=30628 | N=2531 | N=4239 | N=15123 | N=2171 | N=2548 | N=823 | N=1042 | N=1482 | N=85 | N=30 | N=554 |
| Gender (male) | M | 16767 (54.8%) | 1470 (58.1%) | 2473 (58.3%) | 7782 (51.5%) | 1329 (61.2%) | 1693 (66.5%) | 387 (47%) | 575 (55.2%) | 681 (46%) | 57 (67.1%) | 19 (63.3%) | 301 (54.3%) |
|  | F | 13855 (45.2%) | 1061 (41.9%) | 1766 (41.7%) | 7337 (48.5%) | 841 (38.8%) | 854 (33.5%) | 436 (53%) | 467 (44.8%) | 801 (54%) | 28 (32.9%) | 11 (36.7%) | 253 (45.7%) |
| Year of birth |  | 1997<br>[1976;2008] | 2007<br>[1997;2014] | 2005<br>[1995;2012] | 1984<br>[1963;2001] | 2004<br>[1993;2011] | 2002<br>[1991;2010] | 2003<br>[1991;2011] | 2008<br>[1999;2013] | 1991<br>[1972;2006] | 2004<br>[1987;2011] | 2001.5<br>[1995;2008.5] | 1998<br>[1974;2007] |
| Familial case | N | 21854 (76.6%) | 1725 (72.9%) | 3002 (74.1%) | 11949 (85.4%) | 1266 (61.1%) | 1468 (65.4%) | 492 (62.2%) | 771 (76.6%) | 645 (46.5%) | 57 (75%) | 28 (93.3%) | 451 (87.7%) |
|  | Y | 6681 (23.4%) | 641 (27.1%) | 1047 (25.9%) | 2049 (14.6%) | 805 (38.9%) | 778 (34.6%) | 299 (37.8%) | 236 (23.4%) | 742 (53.5%) | 19 (25%) | 2 (6.7%) | 63 (12.3%) |
| Consanguinity | N | 24731 (87.2%) | 1426 (60.8%) | 3247 (82.1%) | 13381 (95.5%) | 1602 (78.8%) | 1699 (75.6%) | 589 (77.3%) | 920 (93.2%) | 1303 (93.1%) | 66 (84.6%) | 28 (100%) | 470 (92.7%) |
|  | Y | 3624 (12.8%) | 921 (39.2%) | 708 (17.9%) | 631 (4.5%) | 432 (21.2%) | 547 (24.4%) | 173 (22.7%) | 67 (6.8%) | 96 (6.9%) | 12 (15.4%) | 0 (0%) | 37 (7.3%) |
| Age at onset of symptoms |  | 3 [0.4;14.1] | 0.3 [0;1.6] | 0.5 [0;2.2] | 8 [2;30] | 2.2 [0.3;7.5] | 0.7 [0.1;3] | 1.2 [0.2;3.8] | 2.8 [0.7;5] | 8 [3;18] | 3 [0.3;8.1] | 2.8 [0.6;4.9] | 3 [0.7;15.4] |
| Immune dysregulation at onset | N | 21482 (77.2%) | 1712 (77.5%) | 3242 (84.1%) | 11175 (80.3%) | 616 (31.6%) | 1944 (86.2%) | 632 (84.9%) | 536 (55%) | 1182 (88.3%) | 49 (67.1%) | 4 (13.3%) | 390 (80.7%) |
|  | Y | 6347 (22.8%) | 498 (22.5%) | 613 (15.9%) | 2741 (19.7%) | 1335 (68.4%) | 311 (13.8%) | 112 (15.1%) | 438 (45%) | 156 (11.7%) | 24 (32.9%) | 26 (86.7%) | 93 (19.3%) |
| Infection at onset | N | 8133 (28.9%) | 436 (19.6%) | 2195 (56.6%) | 1920 (13.6%) | 1161 (59.6%) | 590 (26%) | 108 (14.4%) | 705 (72.5%) | 827 (61.5%) | 44 (60.3%) | 19 (67.9%) | 128 (26.3%) |
|  | Y | 19961 (71.1%) | 1786 (80.4%) | 1680 (43.4%) | 12202 (86.4%) | 787 (40.4%) | 1681 (74%) | 643 (85.6%) | 267 (27.5%) | 518 (38.5%) | 29 (39.7%) | 9 (32.1%) | 359 (73.7%) |
| Malignancy at onset | N | 27294 (98.3%) | 2180 (98.6%) | 3766 (97.8%) | 13620 (98.1%) | 1885 (97.1%) | 2235 (99.4%) | 741 (99.7%) | 969 (99.8%) | 1333 (99.8%) | 69 (94.5%) | 25 (89.3%) | 471 (97.7%) |
|  | Y | 479 (1.7%) | 30 (1.4%) | 85 (2.2%) | 269 (1.9%) | 57 (2.9%) | 13 (0.6%) | 2 (0.3%) | 2 (0.2%) | 3 (0.2%) | 4 (5.5%) | 3 (10.7%) | 11 (2.3%) |
| Syndromic manifestations at onset | N | 24549 (88.3%) | 2046 (92.6%) | 1651 (42.7%) | 13574 (97.7%) | 1798 (92.5%) | 2075 (92.3%) | 683 (91.8%) | 919 (94.6%) | 1274 (95.4%) | 40 (54.8%) | 28 (100%) | 461 (95.4%) |
|  | Y | 3253 (11.7%) | 164 (7.4%) | 2220 (57.3%) | 320 (2.3%) | 145 (7.5%) | 174 (7.7%) | 61 (8.2%) | 52 (5.4%) | 62 (4.6%) | 33 (45.2%) | 0 (0%) | 22 (4.6%) |
| Other onset | N | 24466 (87.9%) | 1992 (90.1%) | 3298 (85.5%) | 12947 (93.1%) | 1782 (91.6%) | 2086 (92.7%) | 685 (92.1%) | 545 (55.6%) | 647 (48.1%) | 59 (80.8%) | 28 (96.6%) | 397 (81.9%) |
|  | Y | 3366 (12.1%) | 220 (9.9%) | 559 (14.5%) | 963 (6.9%) | 163 (8.4%) | 164 (7.3%) | 59 (7.9%) | 436 (44.4%) | 699 (51.9%) | 14 (19.2%) | 1 (3.4%) | 88 (18.1%) |
|  |  | N=30628 | N=2531 | N=4239 | N=15123 | N=2171 | N=2548 | N=823 | N=1042 | N=1482 | N=85 | N=30 | N=554 |
| No clinical symptoms | N | 27605 (94.1%) | 2188 (91%) | 3770 (92.5%) | 13875 (95.9%) | 1926 (91.5%) | 2238 (92.3%) | 735 (91.9%) | 966 (97.2%) | 1326 (91.2%) | 71 (91%) | 28 (100%) | 482 (97.6%) |
|  | Y | 1725 (5.9%) | 217 (9%) | 306 (7.5%) | 595 (4.1%) | 180 (8.5%) | 187 (7.7%) | 65 (8.1%) | 28 (2.8%) | 128 (8.8%) | 7 (9%) | 0 (0%) | 12 (2.4%) |
| Age at clinical diagnosis (CD) |  | 8.2 [2;30.1] | 0.7 [0.3;4.6] | 2.4 [0.4;6.9] | 22 [6.2;44] | 5.1 [1;13] | 2 [0.5;7] | 4.2 [1.1;14] | 5 [2.7;11.4] | 15 [5.2;28.8] | 5 [1.3;13.7] | 5.2 [3;9.8] | 7.5 [2.3;34.1] |
| Delay between onset and CD |  | 1.3 [0.1;5.1] | 0.2 [0;1.3] | 0.8 [0;3.9] | 2.7 [0.7;7.5] | 0.4 [0;3.4] | 0.5 [0.1;2.2] | 1 [0.1;5.6] | 1.5 [0.5;4] | 1.1 [0.1;7] | 0.5 [0.1;2.1] | 0.9 [0.3;3.9] | 1.7 [0.3;4.7] |
| Genetics | Mutation found | 12774 (44.5%) | 1470 (63.1%) | 3647 (88.8%) | 2251 (16%) | 1671 (80.8%) | 1693 (71.8%) | 529 (68.1%) | 662 (65.3%) | 772 (55.6%) | 49 (71%) | 30 (100%) | 0 (0%) |
|  | No mutation found | 2852 (9.9%) | 319 (13.7%) | 79 (1.9%) | 1850 (13.1%) | 195 (9.4%) | 106 (4.5%) | 81 (10.4%) | 71 (7%) | 27 (1.9%) | 9 (13%) | 0 (0%) | 115 (24.7%) |
|  | Not tested | 11956 (41.7%) | 403 (17.3%) | 325 (7.9%) | 9398 (66.8%) | 125 (6%) | 478 (20.3%) | 97 (12.5%) | 250 (24.7%) | 569 (41%) | 8 (11.6%) | 0 (0%) | 303 (65%) |
|  | Pending | 1097 (3.8%) | 136 (5.8%) | 57 (1.4%) | 576 (4.1%) | 76 (3.7%) | 80 (3.4%) | 70 (9%) | 31 (3.1%) | 20 (1.4%) | 3 (4.3%) | 0 (0%) | 48 (10.3%) |
| Age at genetic diagnosis (GD) |  | 5.5 [1.1;15] | 0.9 [0.3;6.7] | 3.2 [0.4;9.7] | 9.4 [2.9;25] | 8.3 [1.8;17.3] | 4.4 [1.1;12.7] | 10 [3.2;20.4] | 8.4 [4;22] | 13.2 [5;26.9] | 8.2 [3.6;17.4] | 9.8 [5;14.1] | NA [NA;NA] |
| Delay between CD and GD |  | 0.3 [0;2.9] | 0.2 [0;1.2] | 0.2 [0;1.7] | 1.3 [0.1;8.8] | 0.2 [0;2.2] | 0.3 [0;2.2] | 0.8 [0.1;5.9] | 0.1 [0;1.1] | 0.2 [0;1.6] | 0.4 [0.1;2.6] | 1 [0.2;4.9] | NA [NA;NA] |
| Reason of GD | Clinical | 8787 (88%) | 975 (87.7%) | 2601 (90.8%) | 1629 (89.2%) | 1192 (82.2%) | 1081 (90.6%) | 380 (85.4%) | 515 (88.6%) | 350 (77.8%) | 39 (92.9%) | 25 (96.2%) | 0 (0%) |
|  | Family | 975 (9.8%) | 77 (6.9%) | 153 (5.3%) | 170 (9.3%) | 245 (16.9%) | 100 (8.4%) | 63 (14.2%) | 65 (11.2%) | 99 (22%) | 3 (7.1%) | 0 (0%) | 0 (0%) |
|  | Neonatal | 114 (1.1%) | 41 (3.7%) | 45 (1.6%) | 20 (1.1%) | 2 (0.1%) | 3 (0.3%) | 0 (0%) | 1 (0.2%) | 1 (0.2%) | 0 (0%) | 1 (3.8%) | 0 (0%) |
|  | Prenatal | 112 (1.1%) | 19 (1.7%) | 64 (2.2%) | 7 (0.4%) | 11 (0.8%) | 9 (0.8%) | 2 (0.4%) | 0 (0%) | 0 (0%) | 0 (0%) | 0 (0%) | 0 (0%) |
| Sequencing method of GD | Gene sequencing | 7693 (83.3%) | 848 (80.1%) | 2122 (82%) | 1421 (83.5%) | 1154 (85.3%) | 970 (87.7%) | 323 (77.5%) | 459 (85.5%) | 347 (84.6%) | 26 (70.3%) | 23 (95.8%) | 0 (0%) |
|  | Non-genetic definitive test | 441 (4.8%) | 35 (3.3%) | 284 (11%) | 11 (0.6%) | 12 (0.9%) | 59 (5.3%) | 3 (0.7%) | 0 (0%) | 36 (8.8%) | 1 (2.7%) | 0 (0%) | 0 (0%) |
|  | Whole exome/genome sequencing | 1100 (11.9%) | 176 (16.6%) | 183 (7.1%) | 270 (15.9%) | 187 (13.8%) | 77 (7%) | 91 (21.8%) | 78 (14.5%) | 27 (6.6%) | 10 (27%) | 1 (4.2%) | 0 (0%) |
| Malignancy | N | 18176 (91.1%) | 1578 (93.4%) | 2491 (88.9%) | 8759 (89.4%) | 1595 (91.4%) | 1609 (95.6%) | 569 (94.7%) | 604 (98.1%) | 632 (97.7%) | 66 (90.4%) | 16 (72.7%) | 257 (89.9%) |
|  | Y | 1783 (8.9%) | 112 (6.6%) | 312 (11.1%) | 1034 (10.6%) | 150 (8.6%) | 74 (4.4%) | 32 (5.3%) | 12 (1.9%) | 15 (2.3%) | 7 (9.6%) | 6 (27.3%) | 29 (10.1%) |
|  |  | N=30628 | N=2531 | N=4239 | N=15123 | N=2171 | N=2548 | N=823 | N=1042 | N=1482 | N=85 | N=30 | N=554 |
| Covid19 | N | 5380 (70.3%) | 456 (81%) | 801 (76.1%) | 2613 (64.1%) | 434 (74.2%) | 363 (81%) | 130 (77.4%) | 260 (74.9%) | 177 (76.3%) | 15 (78.9%) | 6 (75%) | 125 (79.6%) |
|  | Y | 2273 (29.7%) | 107 (19%) | 251 (23.9%) | 1461 (35.9%) | 151 (25.8%) | 85 (19%) | 38 (22.6%) | 87 (25.1%) | 55 (23.7%) | 4 (21.1%) | 2 (25%) | 32 (20.4%) |
| Living status | Alive | 20680 (67.5%) | 1419 (56.1%) | 2580 (60.9%) | 10740 (71%) | 1519 (70%) | 1534 (60.2%) | 546 (66.3%) | 729 (70%) | 1127 (76%) | 52 (61.2%) | 22 (73.3%) | 412 (74.4%) |
|  | Deceased | 3216 (10.5%) | 615 (24.3%) | 676 (15.9%) | 1117 (7.4%) | 356 (16.4%) | 269 (10.6%) | 87 (10.6%) | 23 (2.2%) | 18 (1.2%) | 22 (25.9%) | 3 (10%) | 30 (5.4%) |
|  | Lost to follow-up | 6732 (22%) | 497 (19.6%) | 983 (23.2%) | 3266 (21.6%) | 296 (13.6%) | 745 (29.2%) | 190 (23.1%) | 290 (27.8%) | 337 (22.7%) | 11 (12.9%) | 5 (16.7%) | 112 (20.2%) |
| Age at last follow-up/death |  | 20.2 [9.4;42.6] | 8.7 [2;18.7] | 12.5 [6.3;19.9] | 35.4 [16.9;55.9] | 14.8 [6.5;24.2] | 13.7 [5.8;23.6] | 15 [7;27.3] | 11.2 [6;20.9] | 27.3 [13.6;46.4] | 14.6 [6.3;23.7] | 18.2 [12;22.8] | 19 [8.4;44.1] |
| Living status (stops at CT) | Alive at last follow-up | 24222 (81.1%) | 928 (37.1%) | 3144 (76.5%) | 13522 (92%) | 1313 (61%) | 1680 (67%) | 695 (86.1%) | 956 (95.1%) | 1463 (99.1%) | 41 (49.4%) | 25 (83.3%) | 455 (90.6%) |
|  | Deceased | 2226 (7.5%) | 258 (10.3%) | 496 (12.1%) | 966 (6.6%) | 186 (8.6%) | 181 (7.2%) | 74 (9.2%) | 17 (1.7%) | 11 (0.7%) | 9 (10.8%) | 3 (10%) | 25 (5%) |
|  | Curative therapy | 3431 (11.5%) | 1315 (52.6%) | 470 (11.4%) | 216 (1.5%) | 653 (30.3%) | 648 (25.8%) | 38 (4.7%) | 32 (3.2%) | 2 (0.1%) | 33 (39.8%) | 2 (6.7%) | 22 (4.4%) |
| Age at last follow-up (stops at CT) |  | 19.4 [8;42.6] | 1.3 [0.5;11] | 11.4 [5.2;19.3] | 35.5 [16.8;55.9] | 12.1 [3.2;22.7] | 11.6 [4;21.8] | 14.6 [6.6;27] | 11.1 [5.9;20.7] | 27.4 [13.6;46.5] | 10 [5;20.5] | 18.2 [11;22.8] | 19 [7.8;44.1] |
| Follow-up duration (years) |  | 7.2 [2.6;14] | 4.6 [0.8;12] | 8 [2.9;14] | 7.9 [3.4;14.5] | 5.6 [1.7;11.6] | 7.7 [2;17.2] | 6.6 [1.7;13.1] | 4 [1.6;7.9] | 7.7 [2.1;16.9] | 6.7 [3.4;10.6] | 11.2 [5.2;17.1] | 5.2 [1.5;11] |

Quantitative covariates: Median [Q1;Q3]; qualitative covariates: Effective (Percentage); the order of subgroups from left to right follows the IEI categories I-X of the 2022 IUIS classification available on the end date chosen for inclusion; values Y for “yes/present” for real positive values, N for “no” is shown to reflect the size of the correct comparison group, excluding “unknown” replies; ages in years; CT, curative / definitive therapy; diagnosis via newborn screening is recorded as subcategory of “no clinical symptoms / lab abnormalities only”, where not all choices of the submenu are shown; examples of “non-genetic definitive tests” are FISH, MLPA, or CGH Arrays.

### Representation of genetic IEI diagnoses

Figure 2 shows the absolute numbers of patients with a documented genetic diagnosis (*n*=12,774, 44.5%,) versus those without (*n*=15,905, 55.5%, including patients not genetically tested) by IEI category. As expected, within the largest patient subgroup (PAD), the genetic diagnosis is lacking for the majority of patients (figure 2A) and the clinical diagnosis of CVID is attributed, whereas patients with CID with syndromic features had the highest proportion of genetic diagnoses. The top five genes mutated per IEI category are shown in figure 2B, with *IL2RG* being the most frequently reported germline genetic cause of CID; *22q11.2 deletion syndrome,* of CID with syndromic features; *BTK*, of PAD; *TNFRSF6*, of category IV, diseases of immune dysregulation; *CYBB*, of phagocyte disorders; *STAT1*, of intrinsic or innate immune disorders; *MEFV*, of autoinflammatory syndromes; C1 inhibitor, of complement deficiencies; *RTEL*, of BMF; and somatic *TNFRSF6*, of phenocopies. The top 50 genetic causes of IEI from the ESID-R are shown with patient numbers in descending order in supplementary figure 4.

**Figure 2.**
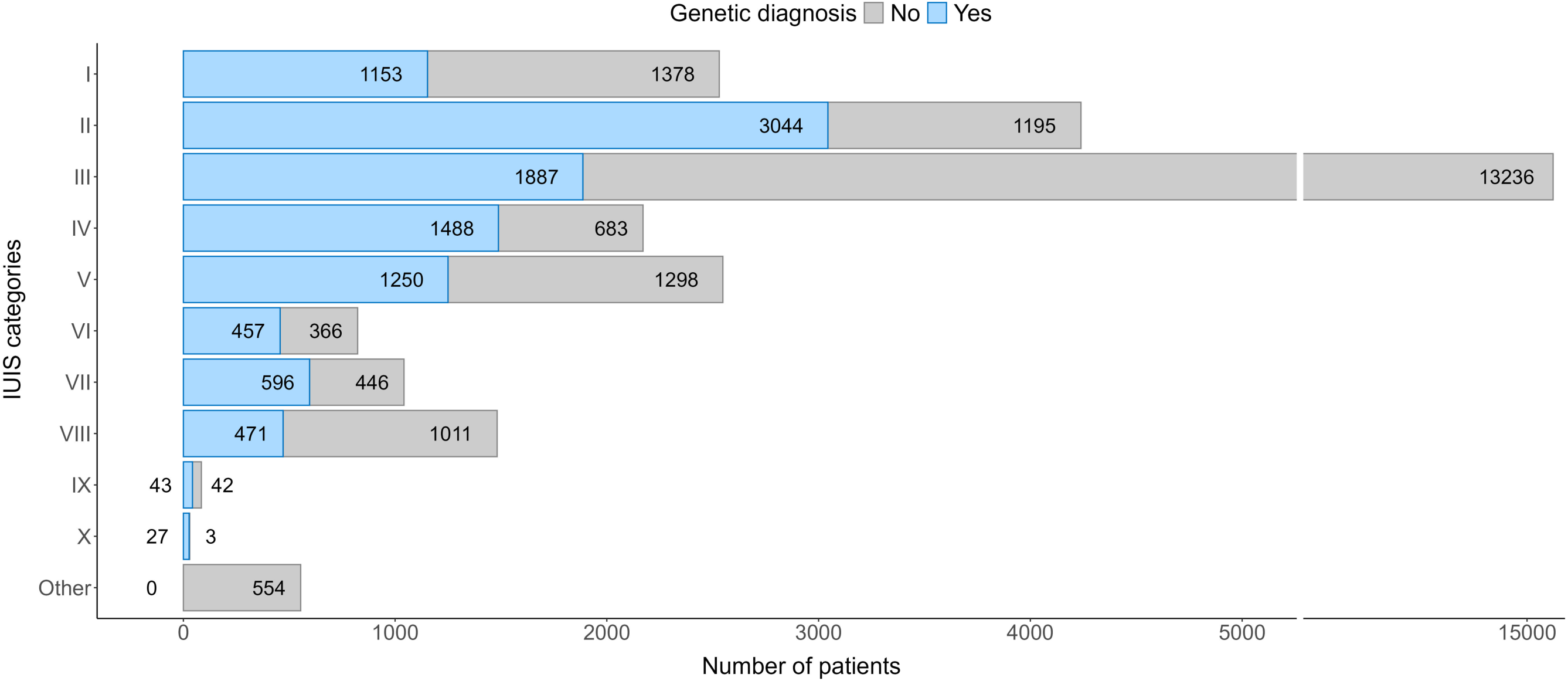
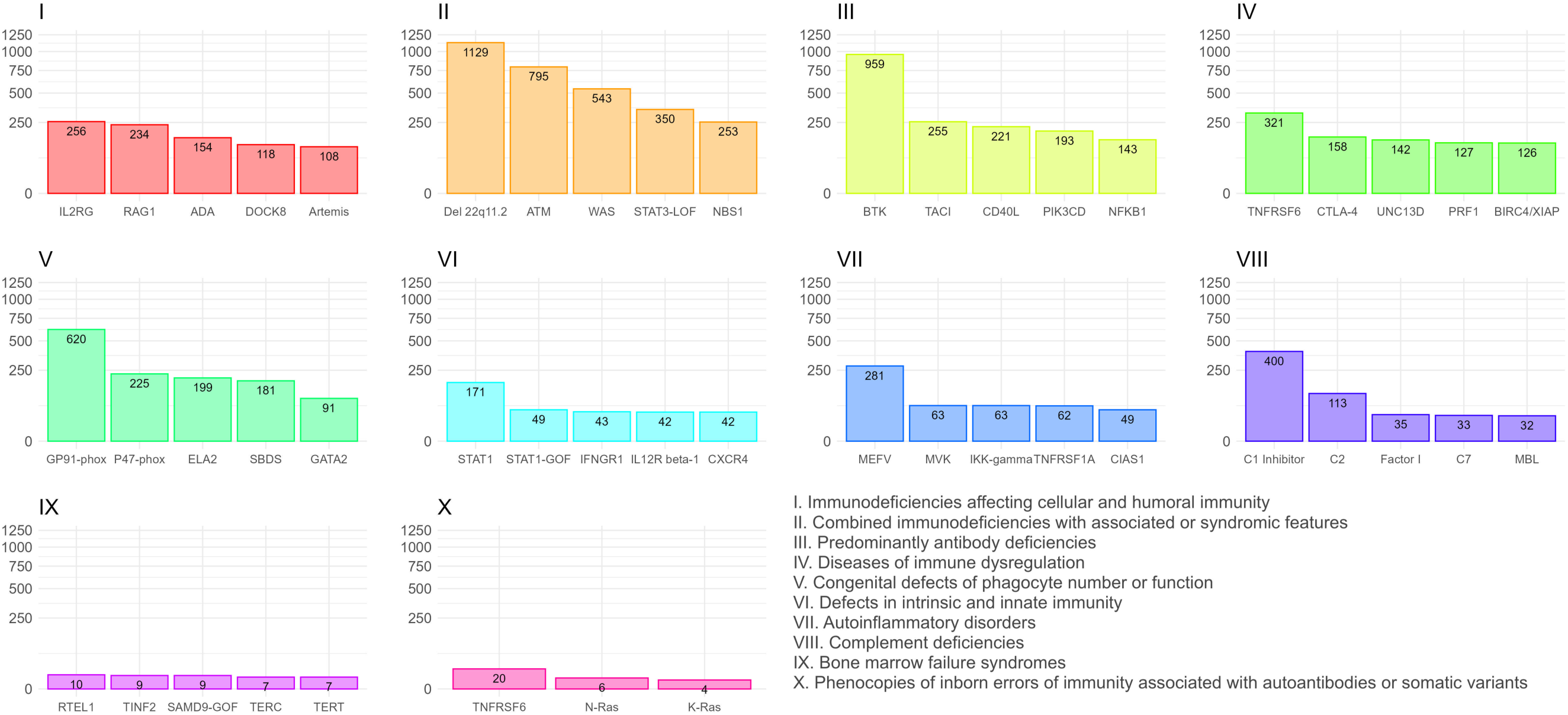
Representation of genetic diagnoses of known IEI/PID in the ESID registry. ***A,*** the number and proportion of patients with a genetic versus those without a genetic diagnosis is shown in descending order. ***B,*** the top 5 genetic defects or deletions registered in the ESIDR per all 10 IUIS categories of IEI are presented; the nomenclature in registry diagnosis and gene fields was not regularly updated/changed, showing, *e.g*., GP91-phox instead of *CYBB* and p47-phox instead of *NCF1*.

### Treatment modalities according to IEI categories

Immunoglobulin replacement therapy (IGRT), hematopoietic stem cell transplantation (HSCT), gene therapy (GT), and splenectomy are recorded in the ESID-R and listed in supplementary table 3. As expected, the highest number and proportion of IGRT-receiving patients is seen in the subgroup of patients with PAD (figure 3). The highest absolute numbers of HSCT procedures were performed in patients with CID, followed by those with PIRD and phagocytic disorders. IGRT and HSCT were documented in patients with IEI/PID in any category. Gene therapy, an evolving curative treatment option devoid of some risks associated with HSCT, such as alloreactivity, was recorded in a descending order for CID, syndromic and phagocyte disorders (figure 3). Splenectomy was recorded relatively frequently in patients with phenocopies, but it was also documented for any IEI category. The use of immune-modulatory treatments such as anti-inflammatory, cell-depleting, or pathway-directed (targeted) therapies and the rare cases of thymus or solid organ transplantation were recorded but are not shown here, as the heterogeneity of the data exceeds the scope of this general report.

**Figure 3.**
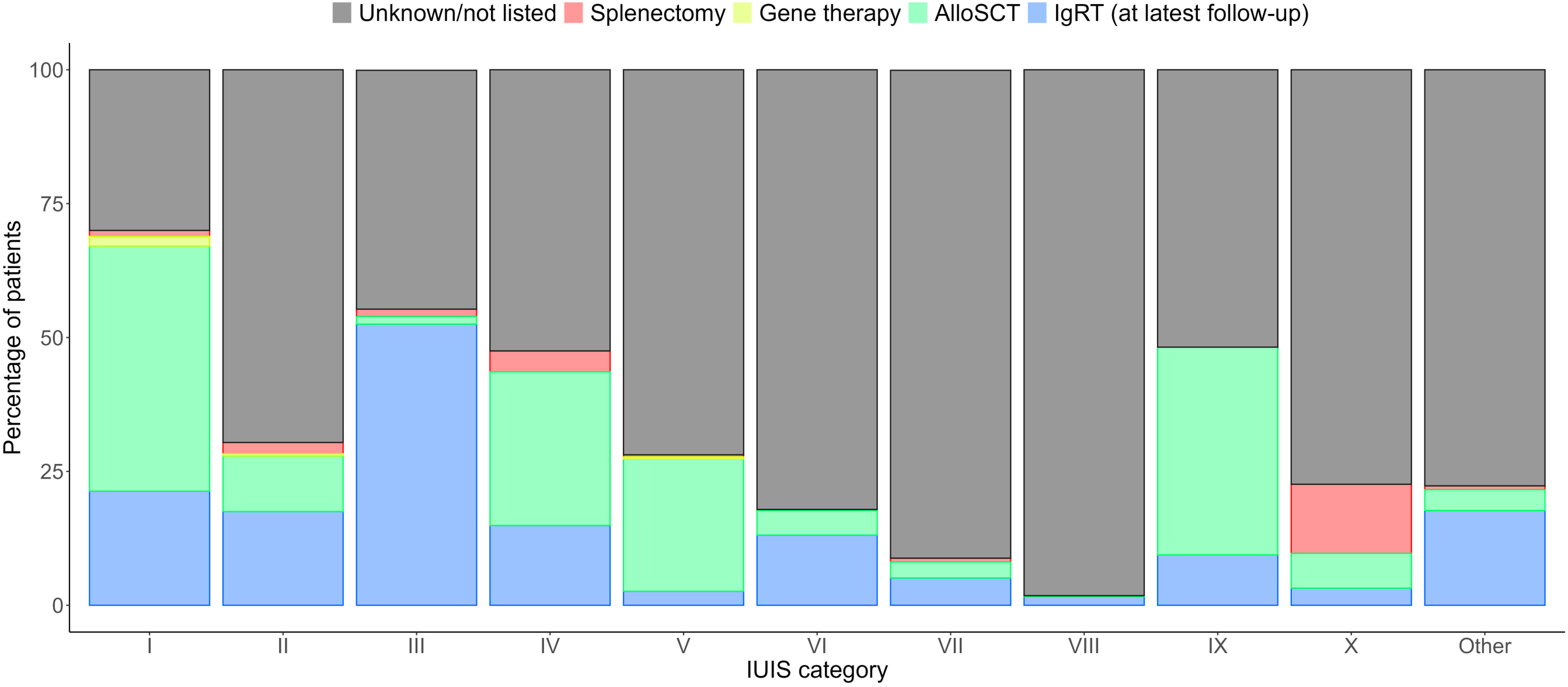
Major treatment modalities of patients with IEI/PID as recorded in the ESID-R. Relative proportions of major treatments such as immunoglobulin replacement therapy (data on route, interval and dose were recorded but are not shown), allogeneic hematopoietic stem cell transplantation (AlloSCT), autologous gene therapy (GT) and on splenectomy. Only a proportion of centers recorded data on immune modifying treatment (not shown), and the ESID-R does not capture data on antimicrobial therapy. “Unknown/not listed” and IgRT refer to the latest follow-up time point.

### Survival probabilities of patients with IEI

We hypothesized that the survival probabilities of patients in the 10 IUIS categories of IEI differed due to their varying predispositions to life-threatening infections, malignancies, autoimmunity, or other manifestations and complications associated with their underlying conditions. Although the data granularity in the entire ESID-R with respect to patient follow-up intervals is not comparable with that in disease-specific prospective cohort studies, we could plot the reversed cumulative incidence function, due to the presence of competing risks, based on the relatively large patient numbers in each IEI category (figure 4; see supplementary figure 5 for the same curves with confidence intervals). Of note, natural biases such as underreporting of patients who died from IEI before diagnosis or of patients with a mild phenotype exist, and numbers at risk increased over the first few years of the observation period (0–97.9 years of age) due to the later time points of inclusion or diagnosis. We detected a steep early decline in survival in many IE categories while curves plateaued, *i*, methodologically, in some where definitive treatments exist, or, *ii*, in genotypes with variable penetrance (*e.g.*, CID, PIRD). Additionally, while patients with PAD showed a continuous decline in survival probability from a young age across all age groups, the decline in the survival of patients with phagocyte and innate immune disorders showed an initial drop. This finding suggests that a proportion of patients are at very high risk during their first five years of life. Diagnoses of early deceased patients with PIRD (n=70 under 5 years of age) were mostly due to disorders with a high risk of hemophagocytic lymphohistiocytosis (HLH; 81.4%); premature deaths in “innate” IEI were frequently due to IRAK4 or MyD88 deficiencies (supplementary table 4). Patients with IEI/PID with syndromic features had a triphasic survival probability. After observing an initial decline in the first four years of life, we detected a second pronounced decline in survival probability in the patients’ 2^nd^ and 3^rd^ decades of life, most likely due to the increased risk of malignancies in many patients in this subgroup (>25%, see supplementary table 2), and a third, relatively steep decline in patients in their 6^th^–7^th^ decades.

**Figure 4.**
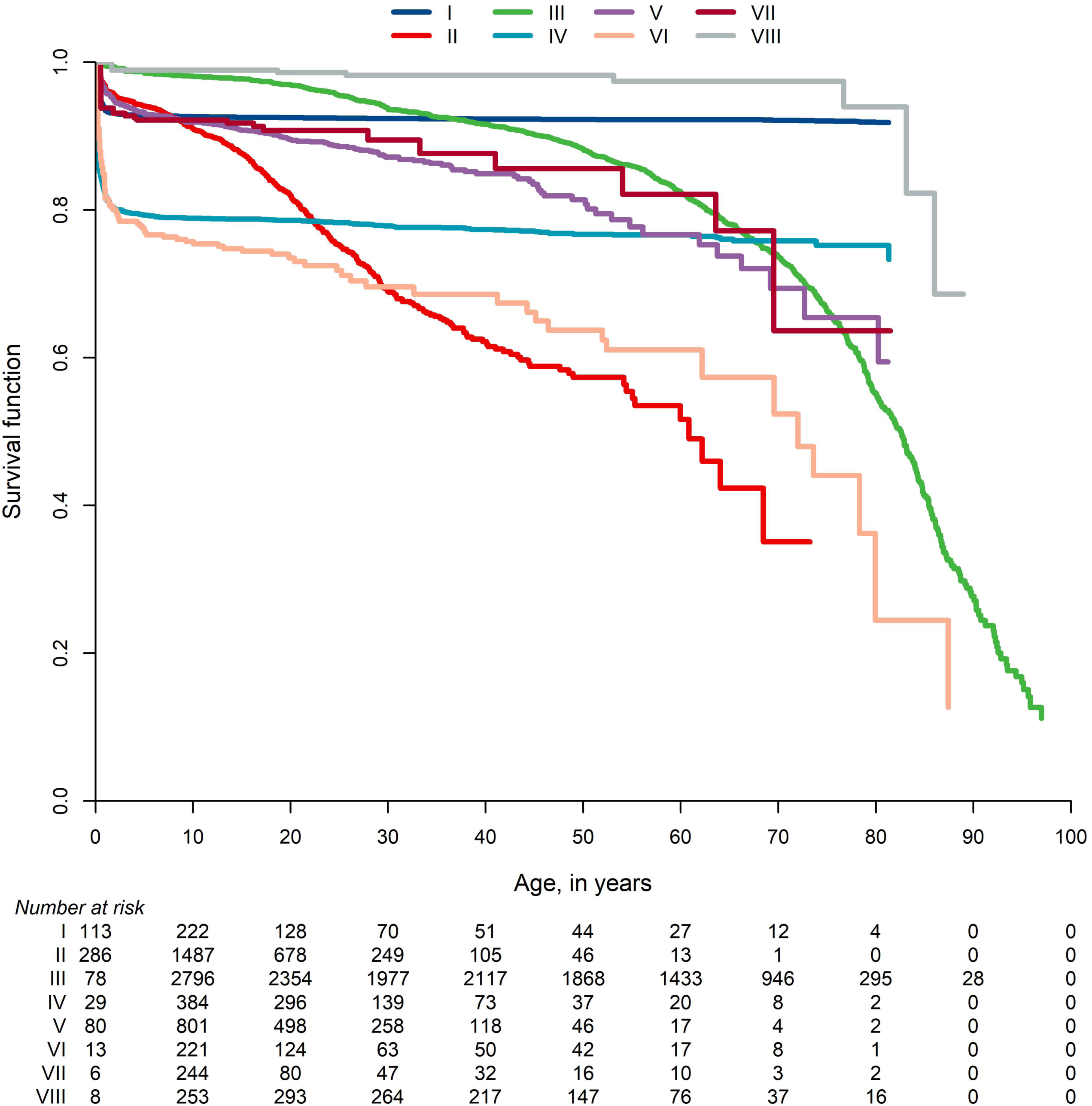
Survival probabilities of main categories of IEI/PID and the living status at last news. ***A,*** Inverse cumulative incidence curves as described and referenced in the supplementary material. Start = age at diagnosis, stop = age at last news, event = living status (0= censored, 1 = deceased first, 2 = curative therapy first). There were 21,206 patients censored, 1,960 patients who deceased first, and 2,901 patients who had a curative therapy first (not showing deaths after curative therapy); roman numbers refer to the IUIS categories for IEI/PID as listed in Figure 2B.

### The ESID-R in the international data source landscape

Other continental or cross-regional IEI/PID registries of varying temporal and geographical depth include the global *Jeffrey Modell Foundation* Centers Network (>94,000 patients), the *USIDNET* registry in the USA (>5,000), the *LASID* registry in Latin America (>9,000), the Australian *AusPIPS* registry (>1,500) as part of the Australasian network, *JSIAD* (>1,200) in Japan as part of the Asian-Pacific (APSID) network, the Canadian registry (*CIEINR*) founded in 2024 and the registries of the *Primary Immune Deficiency Treatment Consortium (PIDTC)* of North America; in addition, many national registries exist inside or outside of ESID(13). Those reported in Europe are shown in supplementary figure 7. Furthermore, the ESID-R is listed as official data source in catalogues of EMA, the European Medical Agency, and of ERDRI, the EU rare disease platform, which are metadata repositories to increase visibility and facilitate the use of rare disease patients’ data.

### Scientific impact of the ESID-R and sub-studies

Up to the end date chosen for data inclusion in this manuscript, 84 peer-reviewed publications resulted from projects deriving data directly from the ESID-R (Figure 5 and ESID website(14)). We evaluated this scientific output by categorizing and counting the publications and their citations as follows: disease-specific natural history studies (*n*=25; 3,658 citations), country-specific epidemiological studies (*n*=19; 2,575 citations), six reviews (422 citations), two on technical aspects (104 citations) and 15 of our own registry-conducted studies (*e.g.*, on first manifestations or on working definitions for the clinical diagnosis of IEI/PID, 1,221 citations), plus 17 studies that could have been assigned to multiple or overlapping categories (figure 5), resulting in a mean citation rate of 95 per publication.

**Figure 5.**
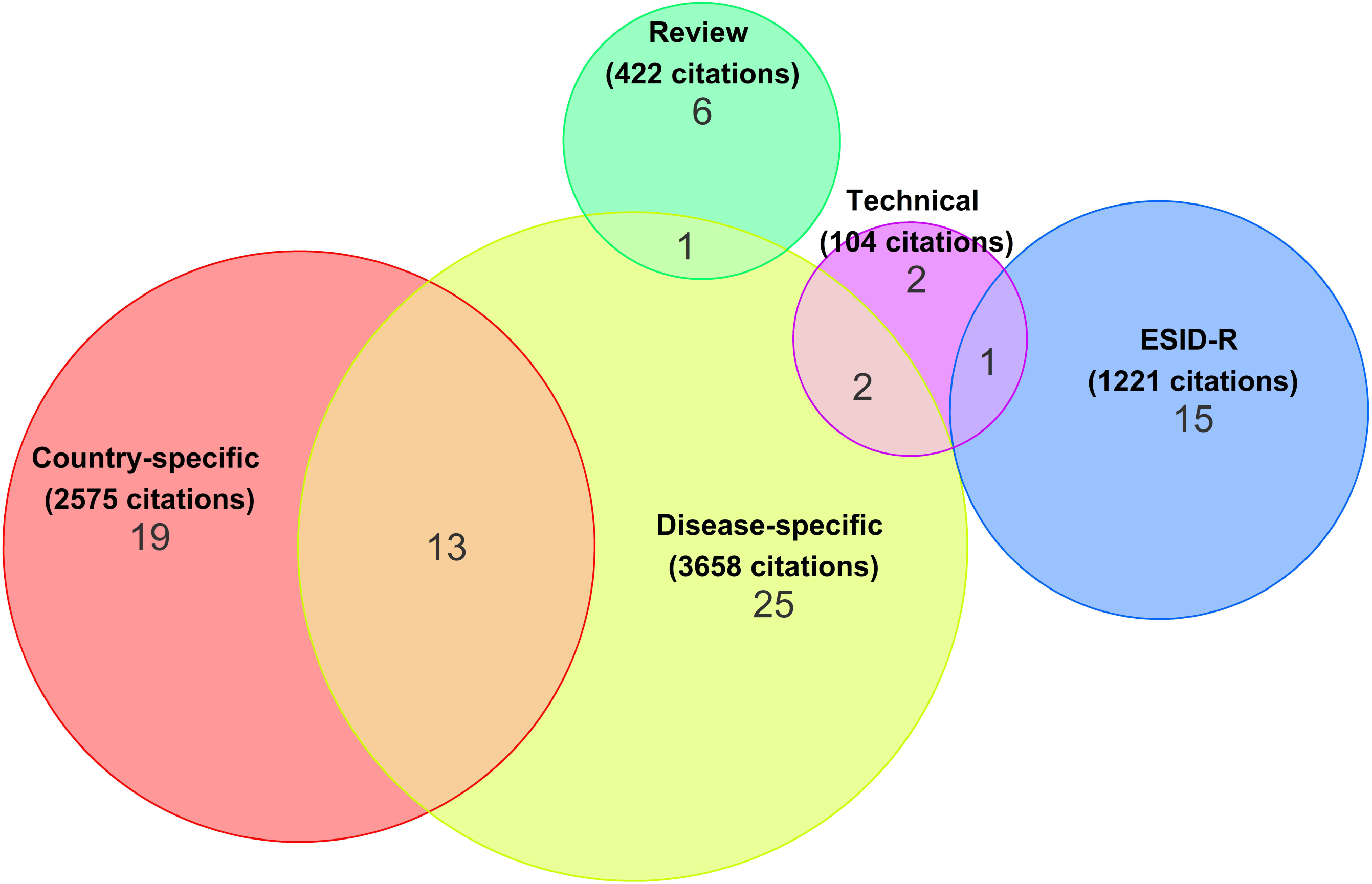
Number, categories and citation counts of ESID-R-based publications. A total of 7,980 citations of 84 peer-reviewed ESID-R-based publications until early 2024 was recorded. See also the website for regular updates of ESID-R-related publications at https://esid.org/working-parties/registry-working-party/registry-publications/

## Discussion

The ESID-R is a large and growing real-world database enabling powerful analyses that have continuously generated epidemiological and disease-specific observational results since 1994. It contributes to knowledge and improvements in patient care in IEI in Europe and beyond. The present analysis illustrates the current distribution of: *i)* IEI diagnoses recorded in contributing centers and countries, *ii)* major treatment modalities between all IEI categories, *iii)*, ages at onset, at clinical and at genetic diagnosis and *iv)* survival probabilities. Our results underline the recommendation to implement newborn screening, potentially even for certain additional IEI other than severe combined immunodeficiencies (SCID). The 7,980 citations of 84 peer-reviewed ESID-R-based publications as of early 2024 demonstrate the success of the ESID-R as platform for research sub-studies and their substantial scientific impact. Hence, the ESID-R provides a clear example of how data collection and collaboration can benefit patients with rare diseases.

### Survival probabilities, early diagnosis, and newborn screening

Newborn screening (NBS) for SCID by measuring T-cell receptor with or without kappa-deleting recombination excision circles in dried blood spots has been implemented in many countries around the world(15-17). A substantial survival benefit for patients diagnosed and treated by early HSCT was demonstrated and recently confirmed in a longitudinal study of the PIDTC(18). The steep early decline in survival we observed in patients with disorders of immune regulation and “innate” IEI may argue for an extension of NBS to other immediately life-threatening IEI (*e.g.*, FHL, XLP1, MyD88/IRAK4 deficiencies and IPEX syndromes). To extend the spectrum of early IEI–NBS, current techniques may be supplemented by RNA sequencing or germline genetic testing, such as targeted sequencing panels, whole exome or genome sequencing. Feasibility studies of such extended NBS are ongoing in regional or national programs(19). However, the vast ethical and social implications of any omics-based addition like baby genome screening will require careful consideration of the risks and benefits(20).

### Methodological and organizational limitations

Physicians and other documentarists regularly collect data for the ESID-R on a voluntary basis, with data quality and quantity (*i.e.*, data depth and accuracy) varying substantially between centers and across countries. Depending on resources dedicated to data documentation, an underestimation of the prevalence of IEI/PID, assumed to be widely similar across Europe, of around 30% was demonstrated(21). While all centers can participate in the ESID-R and scientific sub-studies for free, minimum infrastructure is required. With the rare exception of per-patient fees in pharma-sponsored level-3 studies, ESID does not financially reimburse data entry. The participants mainly benefit from the international academic collaboration and representation, which increases their awareness for disease phenotypes and current clinical research questions. Also, each center may obtain their own (local and regional) epidemiological data within a legally and ethically approved and financially sustainable technical framework (see survey results in the supplementary material). Furthermore, ESID has no means of monitoring the data quality other than by inbuilt checks for logic and completeness at the time point of data entry. So far, automated transfers of data from electronic health records (eHR) into the ESID-R, which have the potential to overcome many of the aforementioned obstacles and to enhance both the quality and quantity of recorded data, have not been attempted due to regulatory obstacles and the heterogeneity of eHRs in the 194 participating centers. All the above, along with the fact that some monogenic IEIs have only been described recently while others have been recognized for decades, introduces a systemic ascertainment bias into the dataset when comparing, *e.g.*, numbers of diagnoses and time of survival. For example, X-linked agammaglobulinemia (caused by mutations in *BTK*)(22) is probably not eight times more prevalent than, *e.g.*, the autosomal-dominant CTLA4-(haplo)insufficiency(23 24) or NFKB1- (haplo)insufficiency(25). In summary, we build on the trust of participants and the success of this huge cooperative effort, to continue the ESID-R in its fourth technical version starting from late 2024, serving as up-to-date simple and free platform for inherently motivated international scientific clinical research collaboration.

### Challenges of reporting genetic data

Collecting data from the heterogenous patient population of individuals with IEI/PID and storing these in a registry presents many challenges. Scientists estimate that current routine next-generation sequencing (NGS) technologies can reveal a molecular cause in approximately 30–35% of IEI cases(19). It is noteworthy that information classifying detected variants by applying ACMG criteria or performing functional validation is not collected in the ESID-R. Phenotype information derived from the ESID-R might support the efforts of the *ClinGen Immunology Gene and Variant Curation Expert Panels*(26). The incorporation of genetic data beyond known pathogenic mutations, ideally linked to precisely defined phenotypes according to the human phenotype ontology (HPO)(27), represents a biologically interesting, future challenge. This includes the verification of variants of unknown significance, intronic changes, somatic mutations and mosaicism, epigenetic factors, variable penetrance and other less-studied factors as non-coding modifier alleles and monoallelic expression(28).

### Link to other registries, synergies, overlapping efforts

From a scientific perspective, it has become increasingly important to compare outcomes for IEI/PID patients with and without cellular therapy (HSCT or gene therapy). The registry of the *European Society for Blood and Marrow Transplantation* (EBMT) currently registers definitive cellular therapies in about 700 patients with IEI per year(29), many of whom are also registered in the ESID registry. Hence, to optimize future research, datasets from the ESID and the EBMT registries for the same patients should be combined. Better alignment of the registries and their data fields is urgently needed to facilitate such studies, and we expect the new technical platform to facilitate data exports and imports alike across providers. Furthermore, the IUIS classification has gradually incorporated more diseases from overlapping specialties (*e.g.,* hematology-oncology, rheumatology, gastroenterology), many of which are (also) covered by other medical societies and patient registries. Thus, although data from many of these patients are stored in the ESID-R, their proportion in the ESID-R does not reflect the real-world distribution. The registry of the *European Reference Network for Rare Immunological and Autoimmune Diseases (ERN-RITA)*, an EU sponsored network of healthcare providers from reference centers for PID/IEI, rheumatology, autoinflammatory and autoimmune diseases, aims to collect common data elements from patients across these disease areas. This initiative may help to avoid redundancy. Finally, using independent (meta-)identifiers such as the *European SPIDER-ID(30)* in all registries would help scientists disentangle the registry landscape.

### Future: AI in registry work for data analysis and interpretation

Advanced software tools and artificial intelligence (AI, *e.g.*, large language models, natural language processing, and machine learning), will certainly transform clinical decision-making and other crucial processes in medicine including the field of IEI.(31 32) Automated eHR scanning and data harvesting processes, ideally for specific terms in accordance with HPO, medical reports, disease classification codes (*e.g.*, ICD-11 or ORPHA codes), but also for free text may be used to identify currently undiagnosed IEI/PID patients. These individuals might benefit from early screening by preventing complications. Accordingly, as a first step, data from an IEI/PID registry like the ESID-R that includes data from patients with an established diagnosis could be used to train AI models and to predict a monogenic IEI or at least the most likely affected pathway in patients who lack a genetic diagnosis. Consequently, patients may undergo screening for disease-specific risks and receive appropriate therapy early in their course. In addition to AI-assisted automated data extraction from eHRs to feed patient registries with structured information, complementing the ESID-R with AI-based tools has the potential to transform this data-collection platform to an electronic IEI/PID patient management assistant in the future.

### Conclusions and Perspectives

The rarity and complexity of many IEI grants them an orphan status regarding pharmaceutical research and development. The feasibility of clinical research, including drug trials and post-authorization efficacy and safety studies, thus depends on large networks of academic institutions and medical specialist societies. As one of the largest registries, containing extensive longitudinal datasets and having a pan-European reach, the ESID-R is likely to remain one of the most relevant scientific registries for patients with IEI.

## Data Availability

All data produced in the present work are contained in the manuscript.

## Acknowledgements

The authors would like to thank all contributors and supporters not already mentioned among the collaborator list of the ESID-R working party (*e.g.*, documentarists, study nurses, data analysts) for their continuous support. Scientific language editing was performed by Dr. Sara Crockett, Graz, Austria (www.saras-science.com). MGS was in part funded by the Styrian Children’s Cancer Aid Foundation (Steirische Kinderkrebshilfe). MHA represents the PID stream of the European Reference Network for Rare Immunological and Autoimmune Diseases (ERN-RITA). CEREDIH thank colleagues from Université de Paris, Imagine Institute, Data Science Platform, INSERM UMR 1163, F-75015, Paris, France for their contribution in the interface between ESID-R and the CEREDIH registry. LDN is supported by the Division of Intramural Research, NIAID, NIH. IM is a senior clinical researcher at FWO Vlaanderen.

## Contributors’ statement

MGS designed the study; MGS and GK wrote the manuscript with contributions from MHA, BG, SG, CS, SE, NM, FC, JLC, LDN, CIES; CS, SS and MP collected data about the global registry landscape; AG performed literature search and analyzed ESID-R-related publication records; MA, MGS and GK analyzed the data and prepared the figures. These and all other authors contributed in varying proportions to the technical, organizational, or structural basis of ESID and the ESID-R and to the collection of patient data. All co-authors contributed to and edited drafts of the original manuscript and figures, and approved the final version.

### List of collaborators (ESID registry working party)

<u>Last name, first name</u>: Abd Elaziz, Dalia; Abdelkader, Sohilla Lofty M.; Abitbol, Avigaelle; Abolhassani, Hassan; Abrahim, Lalash; Abuzakouk, Mohamed; Achir Moussouni, Nabila; Afonso, Veronica; Agyeman, Philipp; Ahlmann, Martina; Aiuti, Alessandro; Akl, Abla; Aksu, Güzide; Albers, Kim; Alecsandru, Meda Diana; Aleinikova, Olga; Aleshkevich, Svetlana; Allende, Luis; Allwood, Zoe; Alsina, Manrique de Lara Laia; Ambrosch-Barsoumian, Daniela; Ameshofer, Lisa; Amour, Kenza; Anadol, Evrim; Ananthachagaran, Ariharan; Andriamanga, Chantal; Andris, Julia; Andritschke, Karin; Angelini, Federica; Ankermann, Tobias; Apel, Katrin; Arami, Siamak; Ardeniz, Ömür; Arkwright, Peter; Arnold, Karina; Ascherl, Rudolf; Assam, Najla; Assia-Batzir, Nurit; Atschekzei, Faranaz; Aumann, Sybille; Aumann, Volker; Aurivillius, Magnus; Ausserer, Bernd; Avcin, Tadej; Aydemir, Sezin; Aygören-Pürsün, Emel; Ayvaz Deniz, Cagdas; Azzari, Chiara; Bach, Perrine; Bachmann, Sophie; Bader, Peter; Bakhtiar, Shahrzad; Bancé, Renate; Bangs, Catherine; Barfusz, Katerina; Baris, Safa; Barlan Isil, B; Bartsch, Michaela; Baumann, Helge; Baumann, Ulrich; Baumeister, Veronika; Baxendale, Helen; Bazen, Suzanne; Beaurain, Beatrice; Beauté, Julien; Bechar-Makhloufi, Mounia; Beck, Norbert; Becker, Brigitta; Becker, Christian; Behrends, Uta; Beider, Renata; Beier, Rita; Belkacem, Amel; Belke, Luisa; Bellert, Sven; Belohradsky, Bernd H.; Ben Slama, Lilia; Ben-Bouzid, Aouatef; Benoît, Vincent; Berdous-Sahed, Thamila; Bergils, Jan; Berglöf, Anna; Bergman, Peter; Bernat-Sitarz, Katarzyna; Bernatoniene, Jolanta; Bernatowska, Ewa; Bernbeck, Benedikt; Bethune, Claire; Beuckmann, Kai; Bhole, Malini; Biegner, Anika-Kerstin; Bielack, Stefan; Bienemann, Kirsten; Bigl, Arndt; Bigorgne, Amélie; Bijl, Marc; Binder, Nadine; Bitzenhofer-Grüber, Michaela; Blanchard Rohner, Geraldine; Blank, Dagmar; Blattmann, Claudia; Blau, Julia; Blaziene, Audra; Blazina, Stefan; Blume, Roswitha; Boardman, Barbara; Bode, Sebastian; Boelens, Jaap-Jan; Boesecke, Christoph; Bogaert, Delfien; Bogner, Johannes; Bohynikova, Nadezda; Booth, Claire; Bordon, Victoria; Borkhardt, Arndt;Börries, Melanie; Borte, Michael; Borte, Stephan; Bossaller, Lukas; Bossard, Madeleine; Botros, Jeannet; Boucherit, Soraya; Boutros, Jeannette; Boyarchuk, Oksana; Boyman, Onur; Boztug, Kaan; Braschler, Thomas; Bravo, Sophie; Bredius, Robbert; Bright, Philip; Brito de Azevedo Amaral, Carolina; Brodszki, Nicholas; Brodt, Grit; Brolund, Allan; Brosselin, Pauline; Brummel, Bastian; Brun-Schmid, Sonja; Brunner, Jürgen; Bruns, Roswitha; Buchta, Christina; Buck, Dietke; Bücker, Aileen; Bührlen, Martina; Burdach, Stefan; Burns, Siobhan; Burton, Janet; Caminal, Luis; Cancrini, Caterina; Canessa, Clementina; Cantoni, Nathan; Capilna, Brindusa; Caracseghi, Fabiola; Caragol, Isabel; Carbone, Javier; Carrabba, Maria; Chamberlain, Latanya;Chandra, Anita; Chantrain, Christophe; Chapel, Helen; Chassot, Julie; Chee, Ronnie; Chinello, Matteo; Chopra, Charu; Chovancova, Zita; Christmann, Martin; Chrzanowska, Krystyna; Ciznar, Peter; Claes, Karlien; Classen, Carl Friedrich; Classen, Martin; Cochino, Alexis-Virgil;Condliffe, Alison M.; Corbacioglu, Selim; Cordeiro, Ana Isabel; Cordier Wynar, Donatienne; Core, Claire; Costes, Laurence; Coulter, Tanya; Courteille, Virginie; Cristina, Maria; Cucuruz, Maria; Dabrowska, Leonik Nel; Dähling, Mandy; Daly, Mary Louise; Daniel, Claudia-Sabrina; Danieli, Maria Giovanna; Darroch, James; Davies, Graham; de Baets, Frans; De Boeck, Christiane; De Gracia Roldan, Javier; de Nadai, Narimene; de Schutter, Iris; De Vergnes, Nathalie; de Vries, Esther; de Witte, Josine; Debert, Theo; Defila, Corina; Deimel, Judith; Delaplace, Diane; Dellepiane, Rosa Maria; Dellert, Nelli; Delliera, Laura; Delor, Anita; Demel, Ulrike; Dempster, John; den Os, M.M.; Dengg, Rosmarie; Desisa, Sora Asfaw; Desta, Alexandra; Detkova, Drahomira; Dewerchin, Maite; Dieli Crimi, Romina; Dilloo, Dagmar; Dimitriou, Florentia; Dinser, Jasmin; Dipani, Nabila; Dirks, Johannes; Dittrich, Anna-Maria; Djermane, Lylia; Dogru, Yagmur; Dogu Esin, Figen; Dombrowski, Angelika; Döring, Michaela; Drabe, Camilla Heldbjerg; Drerup, Susann; Drexel, Barbara; Driessen, G.J.A.; Dückers, Gregor; Dudoit, Yasmine; Duppenthaler, Andrea; Ebetsberger-Dachs, Georg; Ecser, Mate Barnabas; Eekman, Maartje; Ehrat, Rosanna; Eisl, Eva; Ekwall, Olov; El Hawary, Rabab; El-Helou, Sabine M.; El-Marsafy, Aisha; Elbe, Sarina; Elcombe, Suzanne; Eldash, Alia; Eldeen, Youssif Alkady Radwa Salah; Ellerbroek, P.M.; Elling, Roland; Elliott, Jane; Engelhardt, Angelika; Ernst, Diana; Ersoy, Fügen; Esper, Stefanie; Esteves, Isabel; Exley, Andrew; Faber, Martin; Fabio, Giovanna; Fahrni, Gaby; Faletti, Laura Eva;Faletti, Laura Eva; Farber, Claire-Michele; Farela Neves, João; Faria, Emilia; Farkas, Henriette; Farmaki, Evangelia; Faßhauer, Maria; Fasth, Anders; Fätkenheuer, Gerd; Faustmann, Stefanie; Fecker, Gisela; Feighery, Conleth; Feiterna-Sperling, Cornelia; Fernandez-Cruz, Perez Eduardo; Ferreira Concalo, Cordeiro; Ferster, Alice; Feuchtinger, Tobias; Feyen, Oliver; Finke, Daniela; Fitter, Sigrid; Flaschberger, Stefan; Fleckenstein, Lucia; Föll, Dirk; Fontana, Adriano; Forino, Concetta; Förster-Waldl, Elisabeth; Franchet, Nora; Freitag, Dagmar; Frey, Urs P.; Friedel, Elisabeth; Friedrich, Wilhelm; Frisch, Barbara; Frischknecht, Lukas; Fritzemeyer, Stephanie; Gagro, Alenka; Gahr, Manfred; Gambineri, Eleonora; Gamper, Agnes; Gams, Franziska; Ganzow, Astrid; Garcelon, Nicolas; Garcez, Tomaz; Garcia Prat, Marina; Gardulf, Ann; Garibay, Janine; Garwer, Birgit; Gathmann, Jonathan; Gebauer, Corinna; Geberzahn, Linda; Geikowski, Tilman; Geisen, Ulf; Gemander, Christiane; Gennery, Andrew R.; Gerisch, Marie; Gernert, Michael; Gerrer, Katrin; Gerschmann, Stev; Giannini, Carolin; Gil Herrera, Juana; Girndt, Matthias; Girrbach, Ramona; Girschick, Hermann; Gkougkourelas, Ioannis; Gładysz, Dominika; Gnodtke, Elisabeth; Goda, Vera; Goddard, Sarah; Goebel, Daniela; Goffard, Jean-Christophe; Goldacker, Sigune; Gollowitsch, Eva Maria; Gomes, Manuella; Gompels, Mark; González, Míriam; Gonzalez Granado, Luis Ignacio; Gordins, Pavels; Gößling, Katharina; Göschl, Lisa; Gossens, Lucy; Gowin, Ewelina; Graafen, Lea; Graca, Leo; Gradauskiene (Sitkauskiene), Brigita; Graf, Dagmar; Graf, Norbert; Grange, Elliot; Grashoff, H.Anne; Greil, Johann; Grigoriadou, Sofia; Gronlund, Helen; Groß-Wieltsch, Ute; Guerra, Teresa; Guevara-Hoyer, Kissy; Gueye Mor, Seny; Guibert, Noemie; Gülnur, Birgit; Güngör, Tayfun; Guseva, Marina; Guzman, David; Haag, Marcel; Haase, Gabriele; Haenicke, Henriette; Haerynck, Filomeen; Hafsa, Ines; Hagin, David; Hallek, Michael; Hancioglu, Gonca; Handgretinger, Rupert; Hanitsch, Leif G.; Hansen, Susanne; Hariyan, Tetyana; Harrer, Thomas; Hassunah, Pia; Hatzistilianou, Maria; Hauck, Fabian; Hauser, Thomas; Haverkamp, Margje H.; Hayman, Grant; Heath, Paul; Hedrich, Christian; Heeg, Maximilian; Heike, Michael; Heimbrodt, Martin; Heine, Sabine; Heininger, Ulrich; Heinrich, Christian; Heinz, Valerie; Heitger, Andreas; Helbert, Matthew; Helbling, Arthur; Hellige, Antje; Hempel, Julya; Henderson, Karen; Henes, Jörg; Henneke, Philipp; Hennig, Christian; Henrichs, Karin; Herbst, Martin; Hermann, Walter; Hernandez, Manuel; Hernández, Anja; Heropolitanska, Edyta; Herriot, Richard; Herrmann, Friedrich; Herwadkar, Archana; Hess, Christoph; Hess, Ursula; Hesse, Sebastian; Higgins, Sonja; Hilfanova, Anna; Hilpert, Sophie; Hintze, Chantal; Hlaváčková, Eva; Hodl, Isabel; Hodzic, Adna; Hoernes, Miriam; Hoffmann, Christina; Holbro, Andreas; Höllinger, Christiane; Holtsch, Lisa; Holzer, Ursula; Holzinger, Dirk; Hönig, Manfred; Hönscheid, Andrea; Horn, Julia; Horneff, Gerd; Hoyoux, Claire; Hristova, Nataliya; Hübel, Kai; Hübner, Angela; Huemer, Christian; Huissoon, Aarnoud; Hülsmann, Brigitte; Hundsdörfer, Patrick; Huppertz, Hans-Iko; Huß, Kristina; Hussain, Sadia; Husson, Julien; Hüttner-Foehlisch, Tanja; Ijspeert, Hanna; Ikinciogullari, Aydan; İlknur, Kökçü; Irga, Ninela; Jablonka, Alexandra; Jahnz-Rozyk, Karina; Jakob, Marcus; Jakoby, Donate; Jandus, Peter; Jansson, Annette; Jaquet, Melanie; Jardefors, Helene; Jargulinska, Edyta; Jauk, Barbara; Jesenak, Milos; Jilka, Katharina; Jolles, Stephen; Jones, Alison; Jones, Regina; Jonkman-Berk, Birgit; Jönsson, Göran; Joyce, Hilary J.; Juliana, Pricillia; Kabesch, Michael; Kager, Leo; Kahlert, Christian; Kaiser-Labusch, Petra; Kakkas, Ioannis; Kamitz, Dirk; Kanariou, Maria; Kanz, Lothar; Karakoc-Aydiner, Elif; Karanovic, Boris; Kartal-Kaess, Mutlu; Käser, Elisabeth; Katzenstein, Terese L.; Kayserova, Hana; Kelleher, Peter; Kerre, Tessa; Kilic, Sara Sebnem; Kirchner, Martina; Kiwit, Simone; Kiykim, Ayca; Klasen, Jessica; Klaudel-Dreszler, Maja; Klein, Ariane; Klein, Christoph; Klein-Franke, Andreas; Kleine, Ilona; Kleinert, Stefan; Klemann, Christian; Klima, Marion; Kobbe, Robin; Kocacik Uygun, Dilara Fatma; Koch, Melanie; Kochler, Yvonne; Kohistani, Naschla; Kojic, Marina; Kolios, Antonio; Kölsch, Uwe; Koltan, Sylwia; Kondratenko, Irina; Königs, Christoph; Konoplyannikova, Julia; Kopac, Peter; Kopp, Jana; Körholz, Dieter; Körholz, Julia; Korte, Pauline; Kostyuchenko, Larysa; Kötter, Ina; Kracker, Sven; Králícková, Pavlina; Kramm, Christof; Kramme, Philipp;Krausz, Máté; Kreuz, Wolfhart; Kriván, Gergely; Kropshofer, Gabriele; Krüger, Renate; Krystufkova, Olga; Ktistaki, Maria; Kühl, Jörn-Sven; Kühn, Alexander; Kuijpers, Taco W.; Kuis, Wietse; Kullmann, Silke; Kulozik, Andreas; Kumararatne, Dinakantha; Kümmler, Ria; Kündgen, Andrea; Kurenko – Deptuch, Magdalena; Kuss Paula, Cosima; Kütükcüler, Necil; Lafoix-Mignot, Cécile; Lamers, Beate; Lanbeck, Peter; Landais, Paul; Landwehr-Kenzel, Sybille; Langemeyer, Vanessa; Langer, Thorsten; Lankisch, Petra; Lanz, Nadia; Lara, Manrique de; Lara-Villacanas, Eusebia; Laubenthal, Lisa; Laws, Hans-Jürgen; Le Mignot, Loic; Leahy, Ronan; Lee, Jae-Yun; Lehmann, Andrea; Lehmberg, Kai; Lehner, Patricia; Leibfrit, Hans; Leistner, Leoni; Lesch, Petra; Leutner, Simon; Liatsis, Manolis; Libai Véghová, Linda; Liebel, Johanna; Liese, Johannes G.; Linauskiene, Kotryna; Linde, Richard; Linßner, Martina; Lippert, Conrad Ferdinand; Litzman, Jiri; Llobet, Pilar; Lo, Babacar; Lodin, Tariq; Lokaj, Jindrich; Longhino, David; Longhurst, Hilary; Lopes da Silva, Susana; Lorenzen, Catharina; Lougaris, Vassilios; Löw, Doris; Lubatschofski, Annelie; Lucas, Mary; Lutz-Wiegers, Verena; Maaß, Sabine; Maccari, Maria Elena; Macura-Biegun, Anna; Maerz, Vanessa; Maggina, Paraskevi; Mahrenholz, Hannah; Maier, Sarah; Malfroot, Anne; Malinauskiene, Laura; Mannhardt-Laakmann, Wilma; Manson, Ania; Mantkowski, Felicia; Manzey, Petra; Marasco, Carolina; Marcus-Mandelblit, Nufar; Marg, Wolfgang; Markelj, Gasper; Marodi, Laszlo; Marques, José Goncalo; Marques, Laura; Marschall, Karin; Martínez, Natalia; Martinez de la Ossa Saenz-Lopez, Rafael; Martinez-Saguer, Inmaculada; Martire, Baldassarre; Marzollo, Antonio; Masekela, Refiloe; Masjosthusmann, Katja; Masmas, Tania Nicole; Matamoros, Nuria; Mattern, Jutta; Mau-Asam, Pearl; McDermott, Elizabeth; McIntosh, Nichole; Meglic, Karmen Mesko; Meijer, Ruben; Meinhardt, Andrea; Meshaal, Safa; Messaoud, Yasmina; Meyer, Björn; Meyer-Olson, Dirk; Micol, Romain; Micoloc, Bozena; Mielke, Gudrun; Milito, Cinzia; Misbah, Siraj; Mohr, Michael; Mohrmann, Karina; Moin, Mostafa; Molinos, Luis; Möller, Jana; Möller-Nehring, Sarah; Morbach, Henner; Moschese, Viviana; Moser, Olga; Moshous, Despina; Motkowski, Radoslaw; Motwani, Jayashree; Mouftah Galal, Nermeen; Müglich, Carmen; Mukhina, Anna; Muller, Eva; Müller, Christiane; Müller, Gabriele; Müller, Hedi; Müller, Ingo; Müller, Thomas; Müller, Zoe; Müller-Ladner, Ulf; Müller-Stöver, Sarah; Münstermann, Esther; Murtra Garrell, Núria; Muschaweck, Moritz; Mutert, Miriam; Nademi, Zohreh; Naik, Paru; Näke, Andrea; Nalda, Andrea Martin; Nasrullayeva, Gulnara; Naumann-Bartsch, Nora; Nemitz, Verena;Neth, Olaf; Neubauer, Andreas; Neubert, Jennifer; Neumann, Carla; Niehues, Tim; Niemuth, Mara; Nieuwhof, Chris; Nolkemper, Daniela; Noorani, Sadia; Noorlander, Budde Adya; Notheis, Gundula; Nowatsh, Sanam Amelie; Obenga, Gaelle; Ocak, Suheyla; Oker, Mehmet; Olbrich, Peter; Olipra, Anna; Omran, Heymut; Oommen, Prasad; Orosova, Jaroslava; Oskarsdottir, Solveig; Özsahin, Hülya; Pac, Malgorzata; Pachlopnik Schmid, Jana; Pandolfi, Franco; Papadopoulou-Alataki, Efimia; Papatriantafillou-Schmieder, Anna; Parra-Martinez, Alba; Paschenko, Olga; Pašić, Srdjan; Pasnik, Jarek; Patel, Smita; Pavlík, Martin; Paz Artal, Estela; Peeters, Anouk; Pereira da Silva, Sara Branco; Perez-Becker, Ruy; Perlhagen, Markus; Peter, Hans-Hartmut; Pfreundschuh, Michael; Philippet, Pierre; Picard, Capucine; Pietrucha, Barbara; Pietzsch, Leonora; Piquer Gibert, Monica; Pirolt, Kerstin; Plebani, Alessandro; Pleguezuelo, Daniel E.; Pollok, Katrin; Pommerening, Helena; Popihn, Daniela; Poplonek, Aleksandra; Popp, Marina; Porta, Fulvio; Portegys, Jan; Posfay-Barbe, Klara; Potjewijd, Judith; Poulheim, Sebastian; Prader, Seraina; Prämassing-Scherzer, Petra; Prelog, Martina; Prevot, Johan; Price, Arthur; Price, Timothy; Proesmans, Marijke; Provot, Johan; Quinti, Isabella; Raab, Anna; Rack, Anita; Raffac, Stefan; Ramos Oviedo, Eduardo; Randrianomenjanahary, Philippe; Ranohavimparany, Anja; Raptaki, Maria; Rashidzadeh, Roonaka; Rathwallner, Margit; Reda, Shereen; Redouane, Nahida; Regateiro, Frederico S.; Reichenbach, Janine; Reimers, Bianca; Reinhardt, Cornelia; Reinhardt, Dirk; Reinprecht, Anne; Reiß, Tamara; Reisli, Ismail; Renner, Eleonore; Rezaei, Nima; Richter, Alex; Richter, Darko; Rieber, Nikolaus Peter; Rieckehr, Nadja; Riedel, Marion; Riescher, Heidi; Rischewski, Johannes; Ristl, Nicole; Ritterbusch, Henrike; Ritz, Tanja; Rivier, Francois; Rockstroh, Jürgen K.; Roesler, Joachim; Rofiah, Himatur; Rogerson, Elizabeth; Rolfes, Elisabeth; Roller, Beate; Romanyshyn, Yaryna; Rondelli, Roberto; Roosens, Fien; Rösen-Wolff, Angela; Rösler, Valentina; Roth, Johannes; Rothoeft, Tobias; Roubertie, Agathe; Rübsam, Gesa; Rutgers, Abraham; Ryan, Paul; Sach, Gudrun; Sadeghi, Kambis; Sahrbacher, Ulrike; Saidi, Angelika; Sanal Tezcan, Özden; Sanchez-Ramon, Silvia; Santos, Juan Luis; Sargur, Ravishankar; Savchak, Ihor; Savic, Sinisa; Schaaf, Bernhard; Schaefer, Marzena; Schäfe, Christina; Scharbatke, Eva; Schatorje, Ellen; Schauer, Uwe; Scheibenbogen, Carmen; Scheible, Raphael; Scheinecker, Clemens; Schiller, Romana; Schilling, Beatrice; Schilling, Freimut; Schlieben, Steffi; Schmalbach, Thilo; Schmalzing, Marc Thomas; Schmid, Pirmin; Schmidt, Nadine; Schmidt, Reinhold Ernst; Schmitz, Monika; Schneider, Dominik; Schneider, Dominik T.; Schneppenheim, Reinhard; Scholtes, Cathy; Schölvinck, E.H.; Schönberger, Stefan; Schreiber, Stefan; Schrijvers, Rik; Schroll, Andrea; Schruhl, Simone; Schrum, Johanna; Schubert, Ralf; Schuh, Sebastian; Schulz, Ansgar; Schulz, Claudia; Schulze, Ilka; Schulze-Koops, Hendrik; Schulze-Sturm, Ulf; Schumacher, Eva-Maria; Schürmann, Elvira; Schürmann, Gesine; Schuster, Volker; Schwaneck, Eva; Schwarz, Klaus; Schwarz, Tobias; Schwarze-Zander, Carolynne; Schweigerer, Lothar; Schwinger, Wolfgang; Sediva, Anna; Seebach, Jörg; Seger, Reinhard; Segerer, Florian; Selle, Barbara; Seneviratne, Suranjith; Shabanaj, Hatidje; Shcherbina, Anna; Siepelmeyer, Anne; Siepermann, Kathrin; Simon, Anna; Simon, Arne; Simon-Klingenstein, Katja; Simonovic, Marija; Simsen-Baratault, Merlin;Sindram, Elena; Skapenko, Alla; Skarke, Maiken; Skomska-Pawliszak, Malgorzata; Slatter, Mary; Smet, Julie; Sobh, Ali; Sobik, Bettina; Sogkas, Georgios; Sohm, Michael; Solanich-Moreno, Xavier; Soler Palacín, Pere; Sollinger, Franz; Somech, Raz; Sonnenschein, Anja; Soresina, Annarosa; Sornsakrin, Marijke; Soura, Stavrieta; Spaccarotella, Sabrina; Spadaro, Guiseppe; Sparber-Sauer, Monika; Specker, Christof; Speckmann, Carsten; Speidel, Lisa; Speletas, Matthaios; Stachel, Klaus-Daniel; Stadon, Catherine; Stanislas, Aurélie; Stapornwongkul, Cynthia; Staus, Paulina; Steck, Regina; Steele, Cathal; Steffin, Herbert; Steiner, Urs; Steinmann, Sandra; Stevens, Wim; Stiefel, Martina; Stieger, Sarah; Stiehler, Sophie; Stimm, Hermann; Stojanov, Silvia; Stoll, Matthias; Stoppa-Lyonnet, Dominique; Strapatsas, Tobias; Strauß, Gabriele; Streiter, Monika; Strik-Albers, Riet; Strotmann, Gaby; Suárez Casado, Héctor; Subiza, Jose Luis; Sundin, Mikael; Süß, Birgit; Sutter, Fabienne; Szaflarska, Anna; Szemkus, Monika; Tamary, Hannah; Tantou, Sofia; Tarzi, Michael D.; Taschner, Helga; Tedgard, Ulf; Teixeira, Carla; ten Berge, RJM; Tenbrock, Klaus; Tester, Sabine; Tezcan, Ilhan; Thalguter, Sonja; Thalhammer, Julian; Thoma, Katharina; Thomas, Moira; Thomczyk, Fabian; Thon, Vojtech; Thrasher, Adrian; Tierney, Patricia; Tietsch, Nadine; Tommasini, Alberto; Tönnes, Beate; Tony, Hans-Peter; Trachana, Maria; Trapp, Carmen; Tricas, Lourdes; Trindade Neves, Maria Conceicao; Trischler, Jordis; Ubieto, Hugo; Uelzen, Anett; Uhlmann, Annette; Ullrich, Jan; Ullrich, Kurt; Ünal, Ekrem; Urbanski, Gerhard; Urschel, Simon; Uszynska, Aleksandra; Vacca, Angelo; Vaganov, N.N.; Vagedes, Daniel; Valicevic, Stefanie; Vallelian, Florence; van Beem, Rachel T; van Damme, Charlotte; van de Ven, Annick; van den Berg, J. Merlijn; van der Flier, Michiel; van Dissel, J.T.; van Hagen, P.M.; van Montfrans, J.M.; van Ogtrop, Geoffrey; van Rens, Jacqui; van Riel, Christel A.M.P.; van Royen-Kerkhof, Annet; van Well, G.Th.J.; Vasiliki, Antari; Velbri, Sirje; Vencken, Jo; Ventura, Alessandro; Vermeulen, François; Vermylen, Christiane; Viemann, Dorothee; Viereck, Anja; Villa, Anna; Vincke, Jeroen; Vinnemeier-Laubenthal, Lisa; Vo Thi, Kim Duy; Vollbach, Kristina; Volokha, Alla; von Bernuth, Horst; von Bismarck, Philipp; Voss, Rebecca; Voß, Sandra; Vural, Yüksel; Wachuga, Heike; Wagner, Norbert; Wagström, Per; Wahle, Matthias; Wahn, Volker; Wapp, Nadine; Warnatz, Klaus; Warneke, Monika; Warris, Adilia; Wasmuth, Jan-Christian; Wasserfallen, Jean-Blaise; Wawer, Angela; Weber, Manfred; Weemaes, Corrie MR;Wehrle, Julius; Weidinger, Stephan; Weiß, Michael; Weißbarth-Riedel, Elisabeth; Werner, Antje; Wessel, Sara; Westkemper, Marco; Wicher, Monika; Wickmann, Lutz; Wiegert, Sabine; Wiehe, Monique; Wiehler, Katharina; Wiesböck, Lydia; Wiesik-Szewczyk, Ewa; Williams, Anthony; Winkler, Beate; Winkler, Christel; Winkler, Martina; Winkler, Melanie; Wintergerst, Uwe; Wisgrill, Lukas; Witte, Torsten; Wittkowski, Helmut; Wolf, Barbara; Wölke, Sandra; Wolschner, Christina; Wolska-Kusnierz, Beata; Wood, Philip; Workman, Sarita; Worth, Austen; Wortmann, Michaela; Wuillemin, Walter Alfred; Wulffraat, Nico M; Wustrau, Katharina; Wyndham-Thomas, Chloé; Yegin, Olcay; Yildiran, Alisan; Yilmaz, Denise; Young, Patrick; Yucel, Esra; Zečević, Milica; Zepp, Fred; Zetzsche, Klaus; Zeuner, Rainald; Zielen, Stefan; Zimmermann, Martina; Želimir, Erić; Abou-Chahla, Wadih; Aladjidi, Nathalie; Armari-Alla, Corinne; Barlogis, Vincent; Bayart, Sophie; Blanche, Stéphane; Bodet, Damien; Bonnotte, Bernard; Borie, Raphaël; Boutard, Patrick; Boutboul, David; Briandet, Claire; Brion, Jean-Paul; Brouard, Jacques; Castelle, Martin; Cathebras, Pascal; Carausu, Liana; Catherinot, Emilie; Cheikh, Nathalie; Cheminant, Morgane; Comont, Thibault; Couderc, Louis-Jean; Cougoul, Pierre; Deville, Anne; Devoldere, Catherine; Dore, Eric; Dulieu, Fabienne; Durieu, Isabelle; Entz-Werle, Natacha; Fieschi, Claire; Galicier, Lionel; Gandemer, Virginie; Gardembas, Martine; Gourguechon, Clément; Grosbois, Bernard; Guffroy, Aurélien; Guitton, Corinne; Guillerm, Gaëlle; Hamidou, Mohamed; Haro, Sophie; Hatchuel, Yves; Hermine, Olivier; Hoarau, Cyrille; Humbert, Sébastien; Jaccard, Arnaud; Jais, Jean-Philippe; Jannier, Sarah; Jacquot, Serge; Jaussaud, Roland; Jeandel, Pierre-Yves; Jeziorski, Eric; Kebaïli, Kamila; Korganow, Anne-Sophie; Lambotte, Olivier; Lanternier, Fanny; Larroche, Claire; Launay, David; Le Guenno, Guillaume; Le Moigne, Emmanuelle; Lecuit, Marc; Lefèvre, Guillaume; Lelièvre, Jean-Daniel; Levy, Romain; Li-Thiao-Te, Valérie; Lortholary, Olivier; Luca, Luminita; Mallebranche, Coralie; Malphettes, Marion; Marie-Cardine, Aude; Martin-Silva, Nicolas; Masseau, Agathe; Merlin, Etienne; Millot, Frédéric; Miot, Charline; Mirgot, Floriane; Moussouni, Nabila; Mouthon, Luc; Néel, Antoine; Neven, Bénédicte; Nouar, Dalila; Nove-Josserand, Raphaële; Ouachée-Chardin, Marie; Pagnier, Anne; Paillard, Catherine; Pasquet, Marlène; Pellier, Isabelle; Perlat, Antoinette; Piguet, Christophe; Plantaz, Dominique; Rivière, Sophie; Raffray, Loïc; Roblot, Pascal; Rohrlich, Pierre-Simon; Royer, Bruno; Salle, Valéry; Salvator, Hélène; Sarrot-Reynauld, Françoise; Servettaz, Amélie; Stephan, Jean-Louis; Schleinitz, Nicolas; Suarez, Felipe; Swiader, Laure; Taque, Sophie; Thomas, Caroline; Tournilhac, Olivier; Thumerelle, Caroline; Vannier, Jean-Pierre; Viallard, Jean-François.

## Supplementary Material

### Supplementary Text

#### Historical evolution of the ESID registry, objectives, structure, and content

The transition from the first, hardcopy-based, to the first online version of the ESID registry (ESID-R) was headed by Bodo Grimbacher, Freiburg, Germany, with the help of Barbara Frisch and Viviane Knerr as well as programmers Dominik Veit and Stephan Rusch in 2000-2004. The biggest hurdles faced during this first restructuration were ethical considerations, data protection, and obtaining industry funding. The second overhaul was conducted by Stephan Ehl, Freiburg, and programmers Benjamin Gathmann and Stephan Rusch in 2014 and focused on content restructuration. The ESID registry (ESID-R) steering group, led by the elected chairperson of the ESID-R working party, oversees all aspects of the evolution of the registry, including the design, operations, financing, and set-up of dedicated studies.

Specific objectives of the ESID-R include:

1. *Collection of epidemiological data:* The registry contains basic information on patients with IEI, including their medical history, diagnosis, genetics, treatment, and disease progression. Data on the prevalence of these rare diseases and the associated health burden are important for political decision-makers.
2. *Analysis and research:* By analyzing the collected data on these rare diseases through multicenter collaboration, researchers can gain new insights into the pathophysiology and treatment of primary immunodeficiencies.
3. *Support of clinical studies and trials:* Registry data can serve as the basis for clinical studies or trials aimed at developing new therapies, improving the treatment of patients affected by those diseases, and for post-marketing surveillance studies of new drugs and medicinal products.
4. *Improving patient care:* Through the analyses and publication of registry data, the registry helps improve IEI patient care by providing physicians and healthcare providers/decision-makers with access to up-to-date information and practices.

Both versions of the ESID-R operating from 2004-2014 and 2014-2024 had the following key features(1):

1. *Database Architecture:* The ESID-R utilizes a sophisticated database architecture to manage the vast amount of data collected from patients. This database is designed to be scalable, ensuring it can handle the increasing volume of patient records and research findings over time.
2. *Data Security:* Ensuring the security and privacy of patient data is paramount. The ESID-R implements stringent security measures to protect sensitive information, including encryption protocols and access controls
3. *Web-Based Platform:* The ESID-R is accessed through a web-based platform, allowing authorized users to input and access data remotely. This platform is user-friendly and intuitive, facilitating seamless data entry and retrieval processes. Only an up-to-date web-browser and stable internet connection are required to access the system. No additional on-site software needs to be installed/maintained at the documentation centers.
4. *Framework and Technologies:* The ESID-R is built using a combination of programming languages, frameworks, and technologies suited to its specific requirements. Common technologies include SQL databases for data storage, web development frameworks for the user interface, and data analysis tools for reporting and research purposes.

#### The three-level content structure

The old version of the online ESID-R (2004-2014) was closed for documentation on June 25, 2014. During the first main revision of the online registry in 2014, a three-level structure was implemented. Level 1 addresses the task of the registry as a simple epidemiological tool. To achieve patient registration by as many centers as possible, the dataset is kept short and simple such that caregivers who know the patient well can fill in the data without consulting the patient chart in much detail. It contains basic epidemiological information, information about main categories of symptoms at initial presentation and information about some key therapies (immunoglobulins, HSCT, gene therapy). The yearly update is restricted to survival and implementation of major therapies. The Level 1 registration also serves to identify patients for additional in-depth studies including interventional trials. Level 2 was designed for long-term studies on certain diseases or disease groups without a predefined end. Level 2 datasets allow registration of a limited number of clinical manifestations and lab values, that are provided on a yearly basis. It was particularly designed to capture longitudinal data in disease groups, for which the molecular basis has not been fully resolved (such as CVID or specific antibody deficiencies). Again, the datasets are kept simple to limit the burden of registration. Level 3 was designed for studies on particular genetically defined diseases based on a study protocol with a defined endpoint. The level 3 dataset can contain very detailed information including grading of symptoms, more sophisticated lab values, detailed treatment information and quality of life data. Registration intervals are at least yearly. Level 3 studies usually offer financial support for the documenting centers.

The level 2 and 3 projects or ‘sub-studies’ are designed by independent research groups. To apply for implementation of a study in the ESID registry, a statistical evaluation plan, a time and budget forecast, need to be defined in a standardized study proposal, which is assessed for feasibility and to avoid redundancy or competing interests by the registry steering committee and the elected chairperson in regular meetings. There should also be a financial contribution from the PI or research institution to (at least partly) cover the costs associated with the set-up, coordination, data maintenance, and analyses of these registry sub-studies (details on ESID-website, fee structure). When signing the informed consent, patients may choose to have their anonymized data be included either in sub-studies of academic research groups or of pharmaceutical industry-sponsored consortia, or none of them.

### Supplementary Methods

The ESID-R operates based on a technical infrastructure designed to securely receive, store, and enable the analysis of data related to IEI. These data cover, for example, various aspects of the disease, including genetic mutations, clinical symptoms, laboratory test results, imaging findings, and treatment outcomes. Patients with IEI are registered in the web-based database when their disease data are submitted by an authorized physician or healthcare professional or documenting personnel. Centers or sites where patients are cared for are typically university clinics/hospitals and have a data transfer and general data protection regulation (GDPR)-compliant handling agreement and a local IRB approval in place. The data are de-identified and kept strictly confidential to protect patient privacy.

Different measures are applied to avoid duplicates. Firstly, for centers that use the ‘personalised version’ of the registry, where identifying data is kept on a separate server, the pseudonymization tool (Mainzelliste) creates a notification that this (or a similar) patient seems to have been registered before. The system thereby encourages the documenting person to investigate further, before proceeding with the creation of a new patient documentation. This option is not available for centers who enter pseudmonyms directly. Secondly, persons who sign the consent (patients or parents, next of kin) are always asked whether they have signed a similar consent before. These measures reduce the likelihood of double registrations in the first place. Thirdly, for patients who are transferred from pediatric to adult centers, or who move from one region or country to another, usually the caring hospital is aware of the site where the follow-up is done in the future, and, if this is an ESID-R-documenting center, the patient data can be re-assigned to the new center internally, and follow-up for the adult or relocated patient can be continued in the same documentation under the new center’s name.

The Kaplan-Meier curves were generated as described(2). The Kaplan-Meier is done using the start and stop notation to take care of the left truncation effect. Start = age at diagnosis, stop = age at last news, event = living status (0= censored, 1 = deceased first, 2 = curative therapy first). There were 21,206 patients censored, 1,960 patients who deceased first, and 2,901 patients who had a curative therapy first.

### Supplementary Results and Discussion

#### Newborn screening and longitudinal patient documentation in the ESID-R

The ESID-R data structure does not (yet) allow an analysis of improved diagnostic delay and survival in patients diagnosed with SCID in the ESID-R in the NBS (newborn screening) era (supplementary figure 1C). Firstly, NBS for SCID has been performed sequentially over the last decade, but is still not generally available; and secondly, collecting the intended annual follow-up information may cause a system-inherent delay, including underreporting of SCID patients following definitive therapy (or death), before their data are stored in the ESID-R. The ESID-R working party continues to face significant challenges while aiming to integrate data collected over a patient’s entire disease course into a single registry, as this ranges from NBS or clinical diagnosis, through different clinical events, such as a form of definitive therapy, and/or chronic treatment, and/or their optional participation in observational sub-studies and on to a cure or until death. The living status at last news from patient is shown in supplementary figure 6.

#### The ESID-R in the international landscape of registries for patients with rare immunological diseases

To depict the current situation of the global IEI registry landscape and participants’ motivations, we searched PubMed, communicated with representatives of sister societies around the globe, and performed an online survey in 2024. Of 533 publications retrieved by searching for *primary immunodeficiency patient registry* and exclusion of disease-specific or other secondary sub-studies, the remaining publications showed that a vast majority of IEI/PID registries are national epidemiological databases with a limited temporal and geographical scope regarding longitudinal patient data(2-20). They, naturally, contain a substantially smaller number of patient datasets than the continental ESID registry, which also includes data from patients of some neighboring countries. As such, they are valuable and necessary platforms for the connection of healthcare providers and clinical experts, as well as for health politicians and economists. A relatively recently published overview of the global registry landscape and the burden of IEI/PID from 80 countries evaluated the distribution of IEI/PID diagnoses and other factors such as the diagnostic delay or immunoglobulin treatment in relation to the countries’ resources and societal factors such as the rate of consanguinity(21). These data show that the key drivers to implement and maintain an IEI/PID patient registry range from health policy, warranting access to expert care and modern treatments, over patient advocacy, to clinical, biological, and pharmaceutical research and development. Importantly, IPOPI, the global patient organization for IEI, has identified the existence of registries as one of the six key principles of care as part of its PID LIFE Index, a tool to measure the quality of IEI/PID healthcare in a country(22). To analyze the policy of patient inclusion and obtain feedback from documenting physicians or study nurses, we designed a survey distributed through ESID in 2024. A total of 43 physicians from 29 different European and non-European countries participated in this survey (Supplementary Table 5). Seventy percent of respondents (30/43) said they would exclusively register their patients into the ESID-R; 30% of the survey respondents (13/43) said they would also enter patient data into one of the following disease-specific (other than general IEI/PID) registries: *Eurofever, Severe Chronic Neutropenia International Registry (SCNIR), Juvenile Inflammatory Rheumatism (JIR) Cohort,* and the registry of the *EBMT/Center for International Blood and Marrow Transplant Research (CIBMTR)*. Motivations for entering data into the ESID-R included collaborative research (88%), networking (84%), taking part in academic or pharmaceutical studies (84% and 72%), and improvements in patient care (72%) and for IEI education purposes. Reasons given for not entering patient data into the ESID-R were: lack of personnel (84%), legal challenges regarding data transfer (37%), no obvious local benefit (23%), and concerns about small countries being underrepresented (14%). Survey participants stated that they would like a simplified version of the patient consent form, easy interoperability between national registries and the ESID-R, and grant incentives for entering patient data, especially if these were needed at a higher level of granularity.

#### Human Phenotype Ontology (HPO) in IEI/PID, and potential benefit of integrating HPO into the ESID-R

The Human Phenotype Ontology (HPO) comprehensively organizes and defines the phenotypic features of human disease, supporting genomic and phenotypic analyses through semantic similarity and machine learning algorithms(23). Currently, the HPO terminology for IEI/PID still needs to be further developed with help of the community of clinical immunologists and researchers focusing on IEI/PID: *i.e.*, new HPO terms for relevant phenotypic features to be added, and existing HPO terms to be revised, such that the definitions and hierarchical placement in the HPO reflect current knowledge(24). The HPO can be used with the Global Alliance for Genomics and Health (GA4GH) Phenopacket Schema to represent not only phenotypic features, but also measurements, biopsies, and treatments over the entire time course of a disease(25). Collections of phenopackets may support translational research in immunology by providing standardized input data for software programs designed to identify genotype-phenotype correlations, and characterize disease subtypes(26). Integrating the HPO terminology into the ESID-R, would inspire studies within or across IUIS categories, especially in monogenic entities. To make full use of the HPO, better systems for simple but comprehensive and accurate data entry are needed.

### Supplementary Material (Tables)

**Supplementary Table 1.** Contributing countries, sorted by the calculated prevalence per 100,000 inhabitants.

| Country | Registered patients | Genetically diagnosed (%) | Registered patients/100,000 inhabitants | Prevalence /100,000 inhabitants |
| --- | --- | --- | --- | --- |
| Slovakia | 794 | 38.41% | 14.56 | 14.32 |
| France | 9,113 | 48.6% | 13.59 | 10.99 |
| Belgium | 1,333 | 17.78% | 11.61 | 11.36 |
| Switzerland | 658 | 44.68% | 7.67 | 7.55 |
| Slovenia | 129 | 62.79% | 6.18 | 6.18 |
| Germany | 5,056 | 44.96% | 6.08 | 5.73 |
| United Kingdom | 3,949 | 31.22% | 5.91 | 5.51 |
| Ireland | 280 | 39.29% | 5.67 | 5.48 |
| Portugal | 569 | 47.1% | 5.54 | 5.00 |
| Czechia | 573 | 52.88% | 5.37 | 4.98 |
| Netherlands | 715 | 26.85% | 4.13 | 3.96 |
| Greece | 387 | 48.32% | 3.61 | 3.46 |
| Hungary | 315 | 47.94% | 3.22 | 2.68 |
| Austria | 286 | 51.05% | 3.22 | 3.03 |
| Bulgaria | 175 | 38.29% | 2.51 | 2.42 |
| Belarus | 228 | 82.02% | 2.41 | 1.92 |
| Lithuania | 63 | 6.35% | 2.26 | 2.12 |
| Spain | 1,008 | 50.2% | 2.14 | 2.01 |
| Poland | 813 | 66.42% | 2.14 | 1.86 |
| Estonia | 26 | 11.54% | 1.96 | 1.88 |
| Italy | 1,036 | 27.7% | 1.72 | 1.58 |
| Iran | 1,284 | 9.35% | 1.55 | 1.39 |
| Denmark | 77 | 25.97% | 1.32 | 1.31 |
| Serbia | 82 | 34.15% | 1.18 | 1.17 |
| Turkey | 874 | 39.02% | 1.05 | 0.98 |
| Croatia | 36 | 58.33% | 0.89 | 0.79 |
| Egypt | 512 | 43.95% | 0.51 | 0.41 |
| Ukraine | 140 | 82.86% | 0.32 | 0.29 |
| Romania | 50 | 76% | 0.26 | 0.20 |
| Israel | 5 | 80% | 0.06 | 0.06 |
| Russia | 59 | 93.22% | 0.04 | 0.03 |
| Bosnia and Herzegovina | 1 | 100% | 0.03 | 0.03 |
| Sweden | 2 | 100% | 0.02 | 0.02 |

**Supplementary Table 2.**
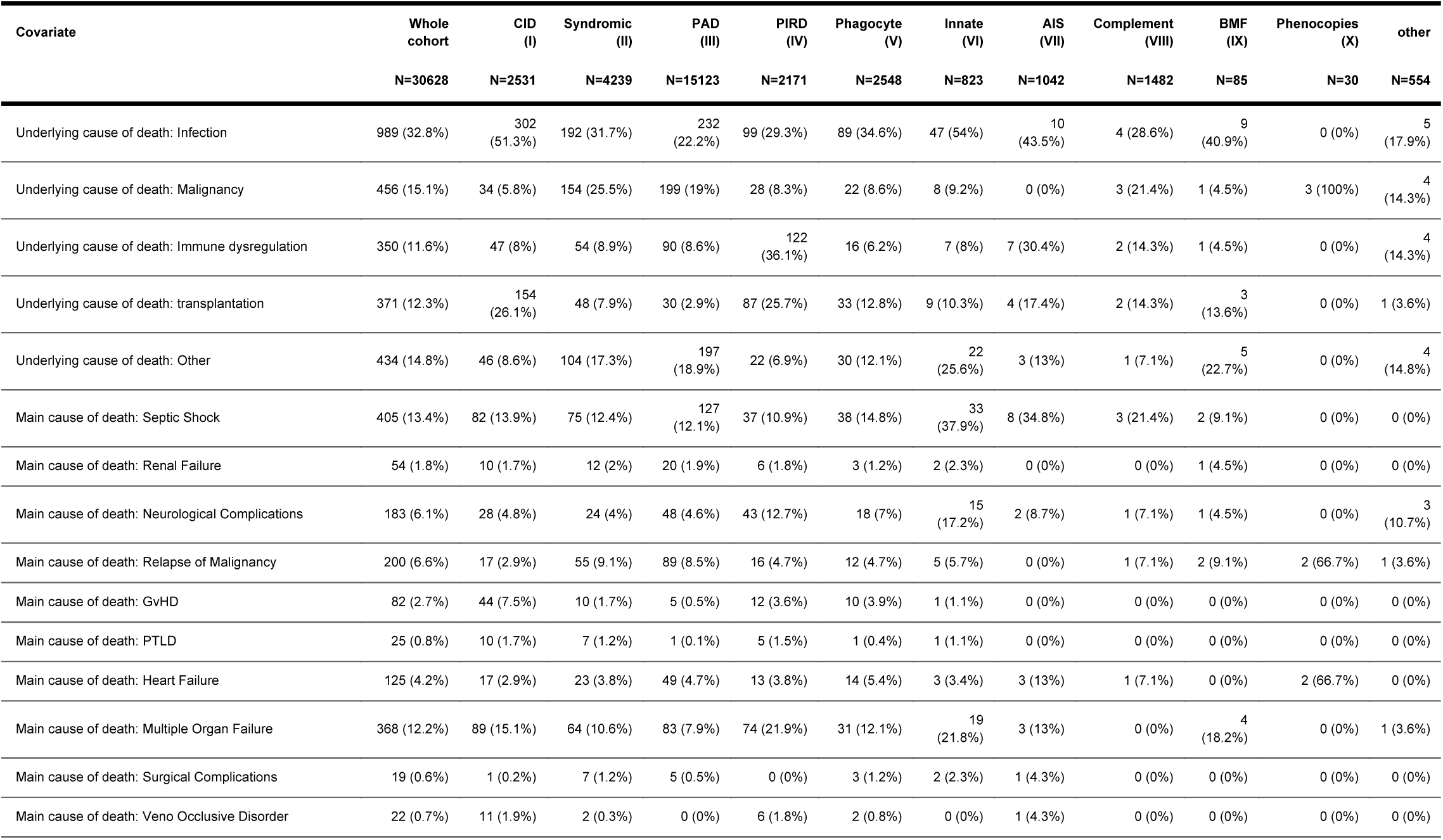
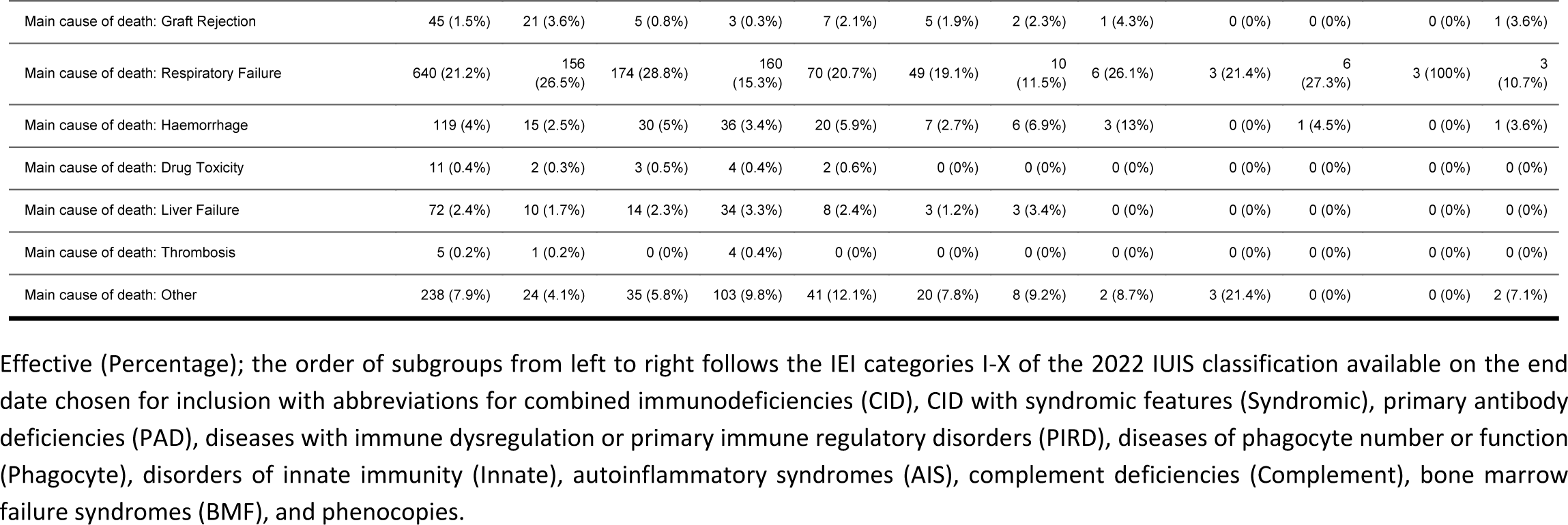
Underlying and main causes of death.

**Supplementary Table 3.** Therapy.

| Covariate | Values | Whole cohort<br>N=30628 | CID (I)<br>N=2531 | Syndromic (II)<br>N=4239 | PAD (III)<br>N=15123 | PIRD (IV)<br>N=2171 | Phagocyte (V)<br>N=2548 | Innate (VI)<br>N=823 | AIS (VII)<br>N=1042 | Complement (VIII)<br>N=1482 | BMF (IX)<br>N=85 | Phenocopies (X)<br>N=30 | other<br>N=554 |
| --- | --- | --- | --- | --- | --- | --- | --- | --- | --- | --- | --- | --- | --- |
| Currently under IgRT | N | 13701 (57.5%) | 1039 (63.8%) | 2280 (74.9%) | 4316 (34.9%) | 1290 (79.2%) | 1864 (96.5%) | 525 (82.9%) | 770 (93.4%) | 1269 (98.2%) | 48 (85.7%) | 20 (95.2%) | 280 (73.9%) |
|  | Y | 10107 (42.5%) | 589 (36.2%) | 764 (25.1%) | 8054 (65.1%) | 339 (20.8%) | 68 (3.5%) | 108 (17.1%) | 54 (6.6%) | 23 (1.8%) | 8 (14.3%) | 1 (4.8%) | 99 (26.1%) |
| IgRT: route | Intravenous | 6410 (51%) | 572 (65.3%) | 600 (58.8%) | 4643 (48.2%) | 324 (63.9%) | 87 (73.1%) | 66 (43.7%) | 41 (57.7%) | 11 (33.3%) | 8 (66.7%) | 2 (66.7%) | 56 (41.2%) |
|  | Subcutaneous | 6119 (48.7%) | 297 (33.9%) | 416 (40.8%) | 4973 (51.6%) | 181 (35.7%) | 32 (26.9%) | 85 (56.3%) | 30 (42.3%) | 21 (63.6%) | 4 (33.3%) | 1 (33.3%) | 79 (58.1%) |
|  | Intramuscular | 17 (0.1%) | 1 (0.1%) | 2 (0.2%) | 12 (0.1%) | 1 (0.2%) | 0 (0%) | 0 (0%) | 0 (0%) | 1 (3%) | 0 (0%) | 0 (0%) | 0 (0%) |
|  | Push (s.c.) | 24 (0.2%) | 6 (0.7%) | 2 (0.2%) | 14 (0.1%) | 1 (0.2%) | 0 (0%) | 0 (0%) | 0 (0%) | 0 (0%) | 0 (0%) | 0 (0%) | 1 (0.7%) |
| Age at first IgRT |  | 19.3 [5.2;41.8] | 2 [0.5;10.3] | 5 [1.3;11.5] | 28.6 [8.7;46.7] | 10.2 [4.2;19.1] | 6.3 [2.1;11.8] | 6.7 [2.3;13.3] | 5.6 [1.9;15.9] | 9.7 [5.6;18] | 1.6 [0.7;7.3] | 7 [6.1;14] | 17.1 [5.9;45.2] |
| AlloSCT | N | 27263 (89%) | 1261 (49.8%) | 3783 (89.2%) | 14907 (98.6%) | 1518 (69.9%) | 1907 (74.8%) | 785 (95.4%) | 1010 (96.9%) | 1480 (99.9%) | 52 (61.2%) | 28 (93.3%) | 532 (96%) |
|  | Y | 3365 (11%) | 1270 (50.2%) | 456 (10.8%) | 216 (1.4%) | 653 (30.1%) | 641 (25.2%) | 38 (4.6%) | 32 (3.1%) | 2 (0.1%) | 33 (38.8%) | 2 (6.7%) | 22 (4%) |
| Age at first AlloSCT |  | 2 [0.6;8.1] | 0.7 [0.4;1.7] | 2.3 [1.1;6.5] | 9.5 [3.5;17.7] | 2.4 [0.5;8.9] | 6.1 [2.2;13.1] | 7.8 [3.3;10.7] | 4 [1.5;7] | 11.4 [10.5;12.3] | 10 [5.1;16] | 7.9 [6.6;9.2] | 7.4 [4.9;14.4] |
| Gene therapy | N | 29070 (99.7%) | 2327 (97.9%) | 4011 (99.6%) | 14465 (100%) | 2086 (100%) | 2452 (99.6%) | 793 (100%) | 923 (100%) | 1411 (100%) | 81 (100%) | 30 (100%) | 491 (100%) |
|  | Y | 77 (0.3%) | 50 (2.1%) | 17 (0.4%) | 0 (0%) | 0 (0%) | 10 (0.4%) | 0 (0%) | 0 (0%) | 0 (0%) | 0 (0%) | 0 (0%) | 0 (0%) |
| Age at first gene therapy |  | 2.3 [0.8;4.6] | 0.9 [0.6;2.9] | 3.6 [2.8;7.4] | NA [NA;NA] | NA [NA;NA] | 15.1 [7.7;22.9] | NA [NA;NA] | NA [NA;NA] | NA [NA;NA] | NA [NA;NA] | NA [NA;NA] | NA [NA;NA] |
| Splenectomy | N | 19732 | 1565 | 2681 (96.8%) | 9846 | 1620 | 1485 | 603 | 753 (99.1%) | 789 (99.9%) | 55 | 21 (84%) | 314 (98.7%) |

| Covariate | Values | Whole cohort | CID (I) | Syndromic (II) | PAD (III) | PIRD (IV) | Phagocyte (V) | Innate (VI) | AIS (VII) | Complement (VIII) | BMF (IX) | Phenocopies (X) | other |
| --- | --- | --- | --- | --- | --- | --- | --- | --- | --- | --- | --- | --- | --- |
|  |  | (97.8%) | (98.1%) |  | (97.9%) | (94.8%) | (99.3%) | (99.7%) |  |  | (100%) |  |  |
|  | Y | 453 (2.2%) | 30 (1.9%) | 90 (3.2%) | 216 (2.1%) | 89 (5.2%) | 10 (0.7%) | 2 (0.3%) | 7 (0.9%) | 1 (0.1%) | 0 (0%) | 4 (16%) | 4 (1.3%) |
| Age at splenectomy |  | 17.9 [7.3;33.3] | 16 [9;24] | 5.4 [2.3;8] | 31.3 [20.1;42] | 11.3 [6.7;20.2] | 8.9 [4.5;21.3] | 2.8 [2.8;2.8] | 21.2 [17.9;36.1] | 33.9 [33.9;33.9] | NA [NA;NA] | 7.2 [4.6;8.6] | 19.3 [15.7;25.3] |

Quantitative covariates: Median [Q1;Q3]; qualitative covariates: Effective (Percentage)

**Supplementary Table 4.**
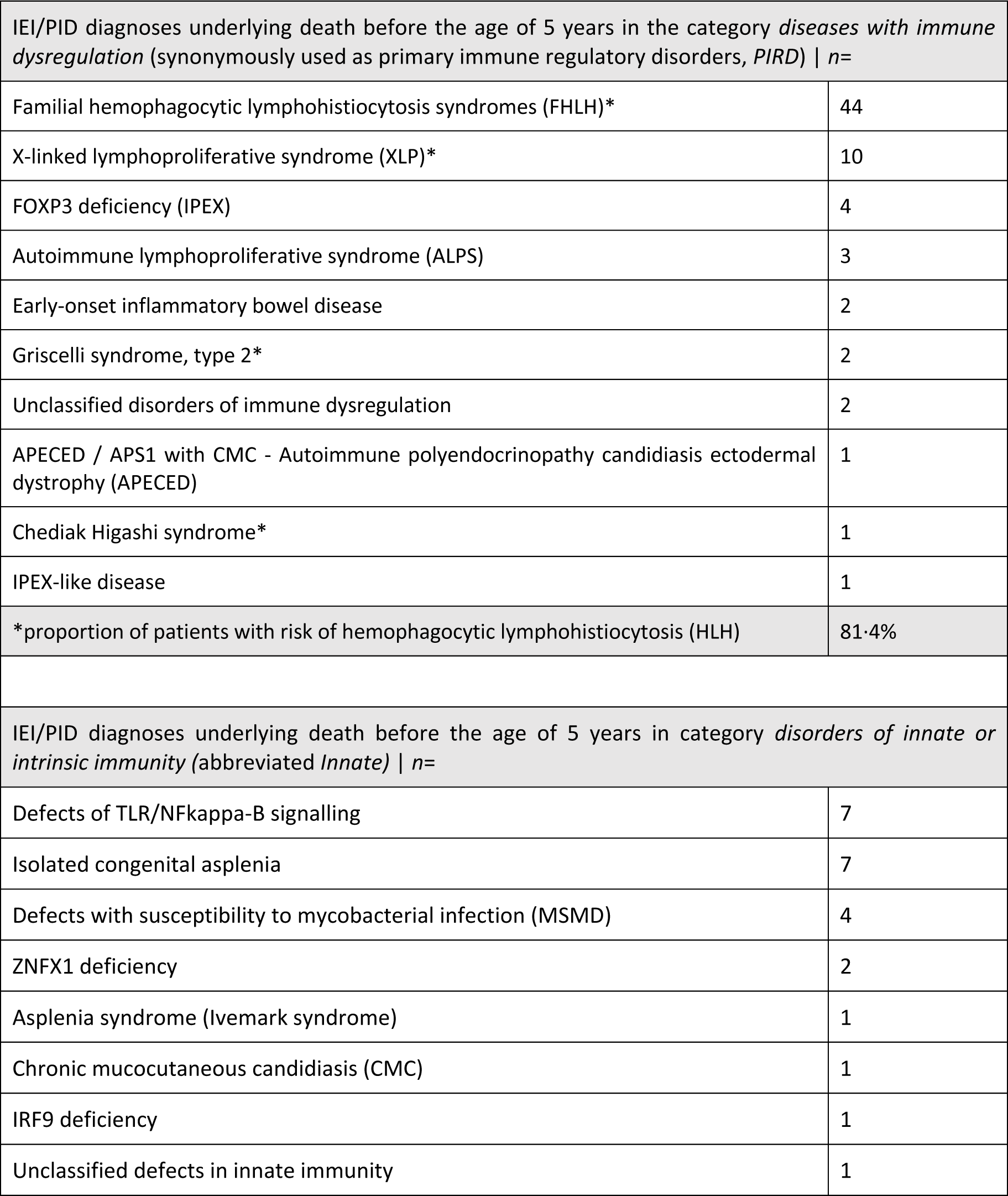
Sub-analysis of IEI/PID diagnoses in patients who died before the age of 5 years in two subcategories of IEI/PID.

**Supplementary Table 5.** Institutions of participants of the 2024 ESID-R survey on the international IEI/PID registry landscape.

| **Western Europe** | **Eastern Europe** | **Other countries** |
| --- | --- | --- |
| Denmark (Copenhagen) | Armenia (Yerevan) | Australia (Melbourne) |
| Finland (Helsinki) | Belarus (Minsk) | Brazil (São Paulo) |
| Germany<br>(Dresden, Freiburg, Leipzig, Munich) | Bulgaria (Sofia) | Canada (Halifax) |
| Ireland (Dublin) | Czech Republic (Prague) | Egypt (Cairo) |
| Italy (Milan) | Georgia (Tbilisi) | Iran (Teheran) |
| Portugal (Coimbra) | Hungary (Budapest) | Sudan (Khartoum) |
| Spain (Barcelona) | Latvia (Riga) | Turkey (Istanbul) |
| Sweden (Jönköping) | Romania (Bucharest) | USA (Philadelphia) |
| Switzerland (Geneva, Zurich) | Russia (Moscow) |  |
| United Kingdom (Sheffield) | Serbia (Belgrade) |  |
|  | Ukraine (Kyiv) |  |

*The survey participants chose to reply (or not) to a survey which had been distributed via the ESID-R* *listserv and was also posted on the website during the summer of 2024.*

### Supplementary Material (Figures)

**Supplementary Figure 1A.**
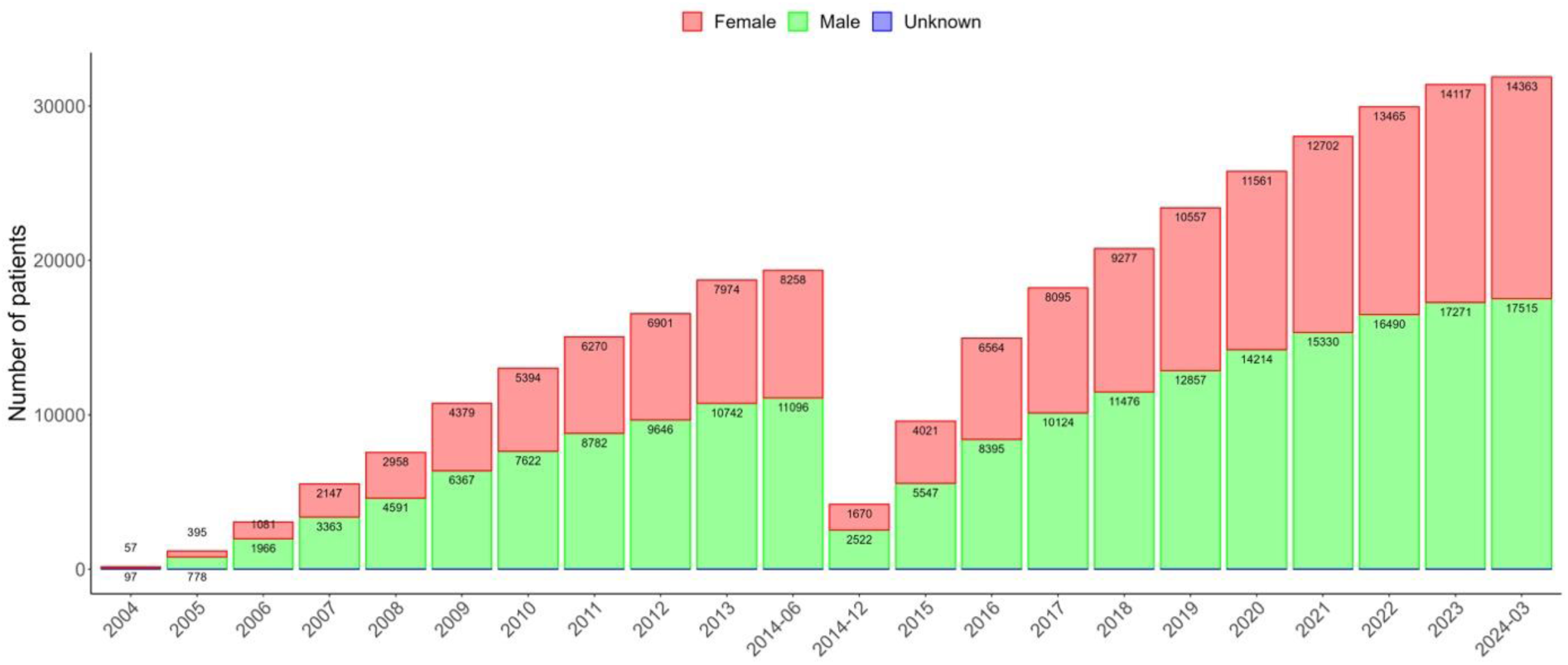
Cumulative number of patients registered in the ESID-R. The total number shown here is 31,889 patients (male *n*=17,515; female *n*=14,363; unknown *n*=11), before exclusion of those discharged (*n*=538), those with secondary immunodeficiency (*n*=417) and those without a definitive diagnosis of IEI/PID (*n*=306). Data shown for the **online** Registry only. Data from the very first version (hardcopy) was not transferred to the first online registry, but newly entered. According to “The Source https://esid.org/wp-content/uploads/2024/03/ESID_TheSource_2003.pdf” the total number in 2003 was 9707 pts.

**Supplementary Figure 1B.**
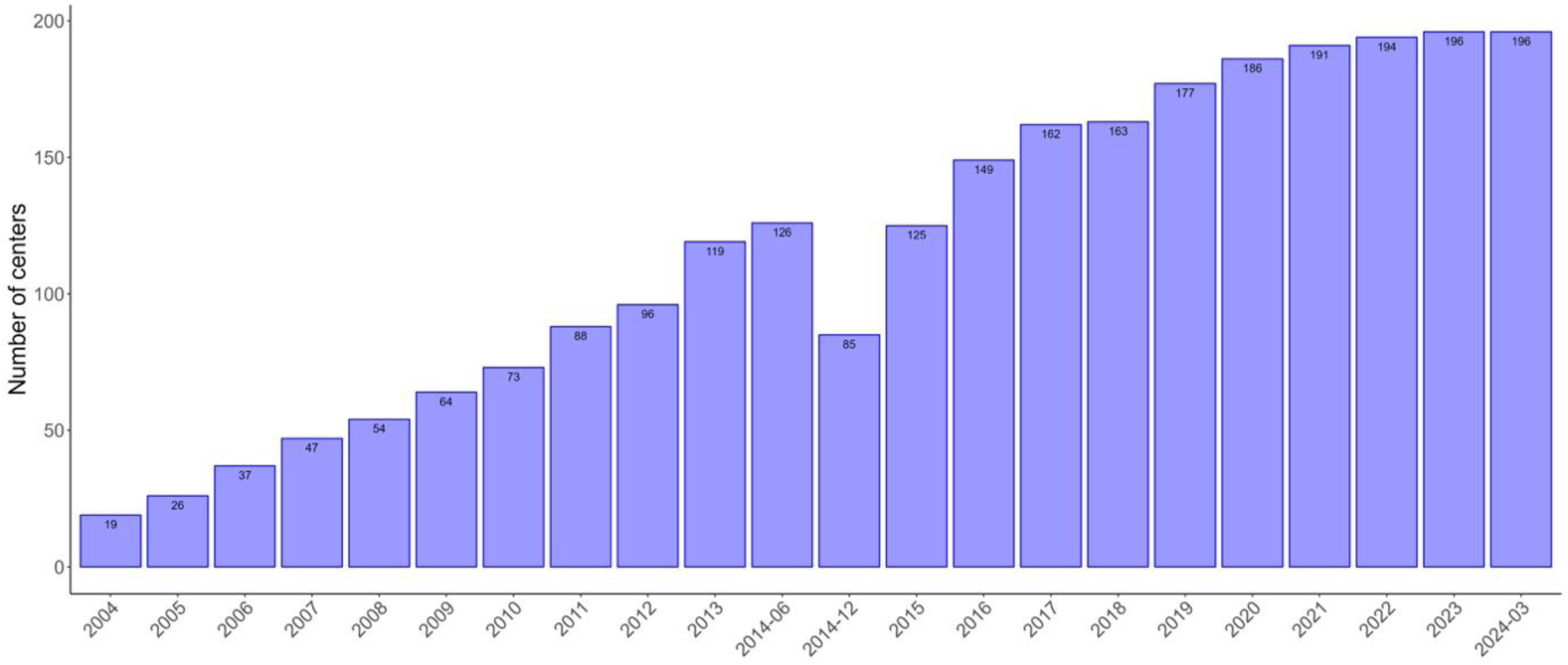
Cumulative number of ESID-R participating centers.

**Supplementary Figure 1C.**
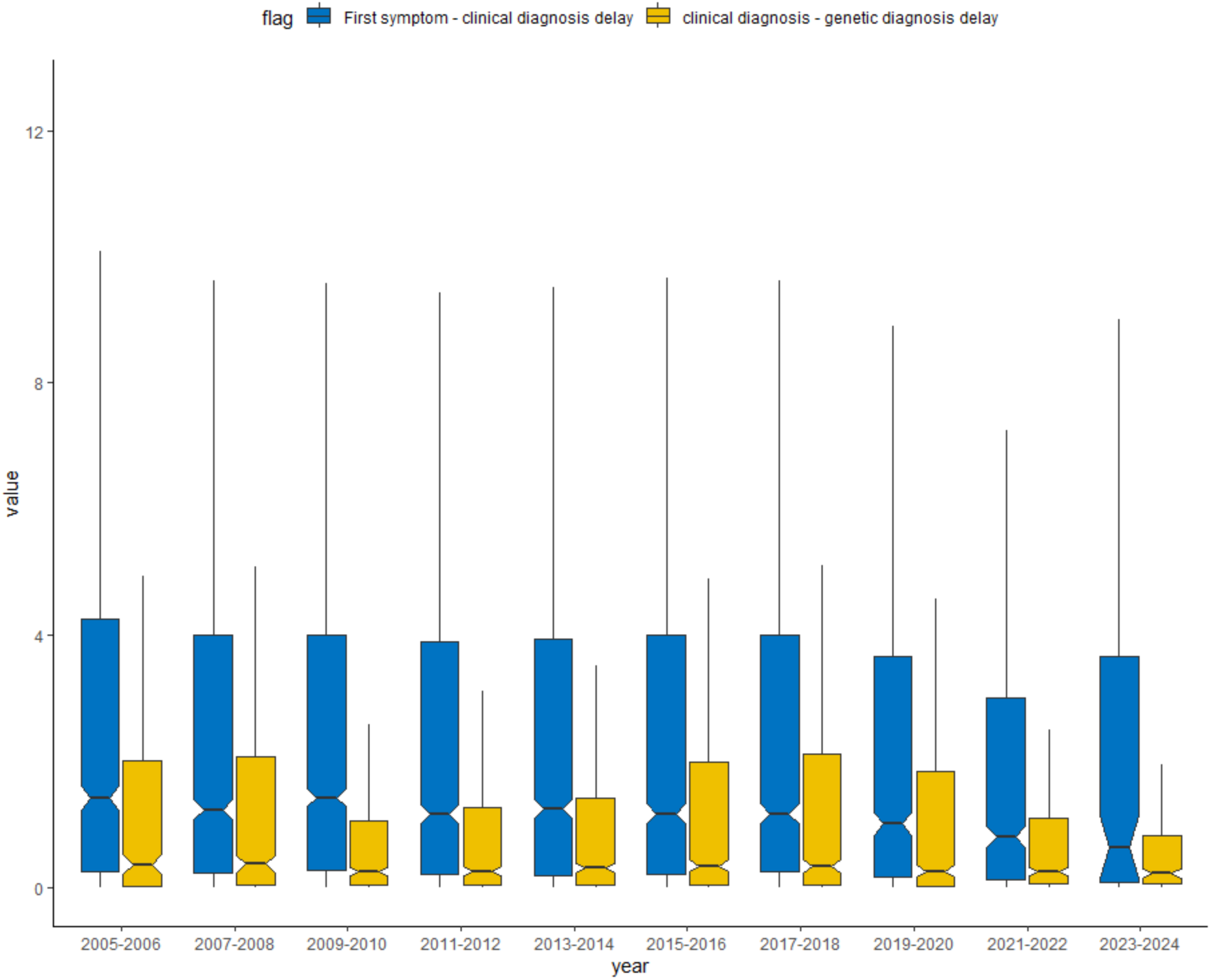
Diagnostic delay (in years from first manifestation to clinical diagnosis and from clinical diagnosis to genetic diagnosis) over the last 20 years of the ESID-R. The y-axis shows the year of clinical/genetic diagnosis.

**Supplementary Figure 2.**
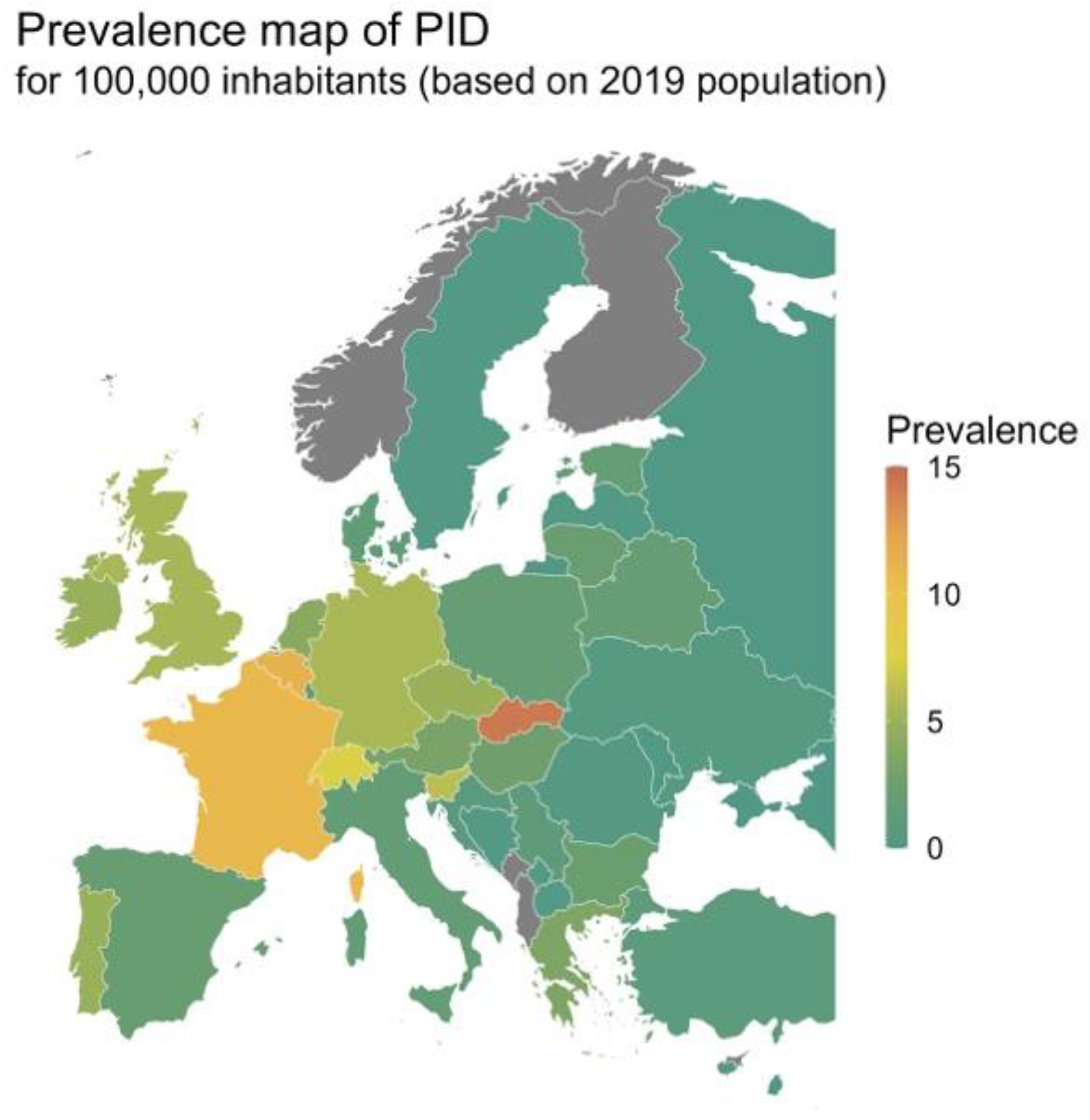
IEI/PID minimal prevalence map according to ESID-R patient numbers and the 2019 population (same data as in Supplementary Table 1).

**Supplementary Figure 3.**
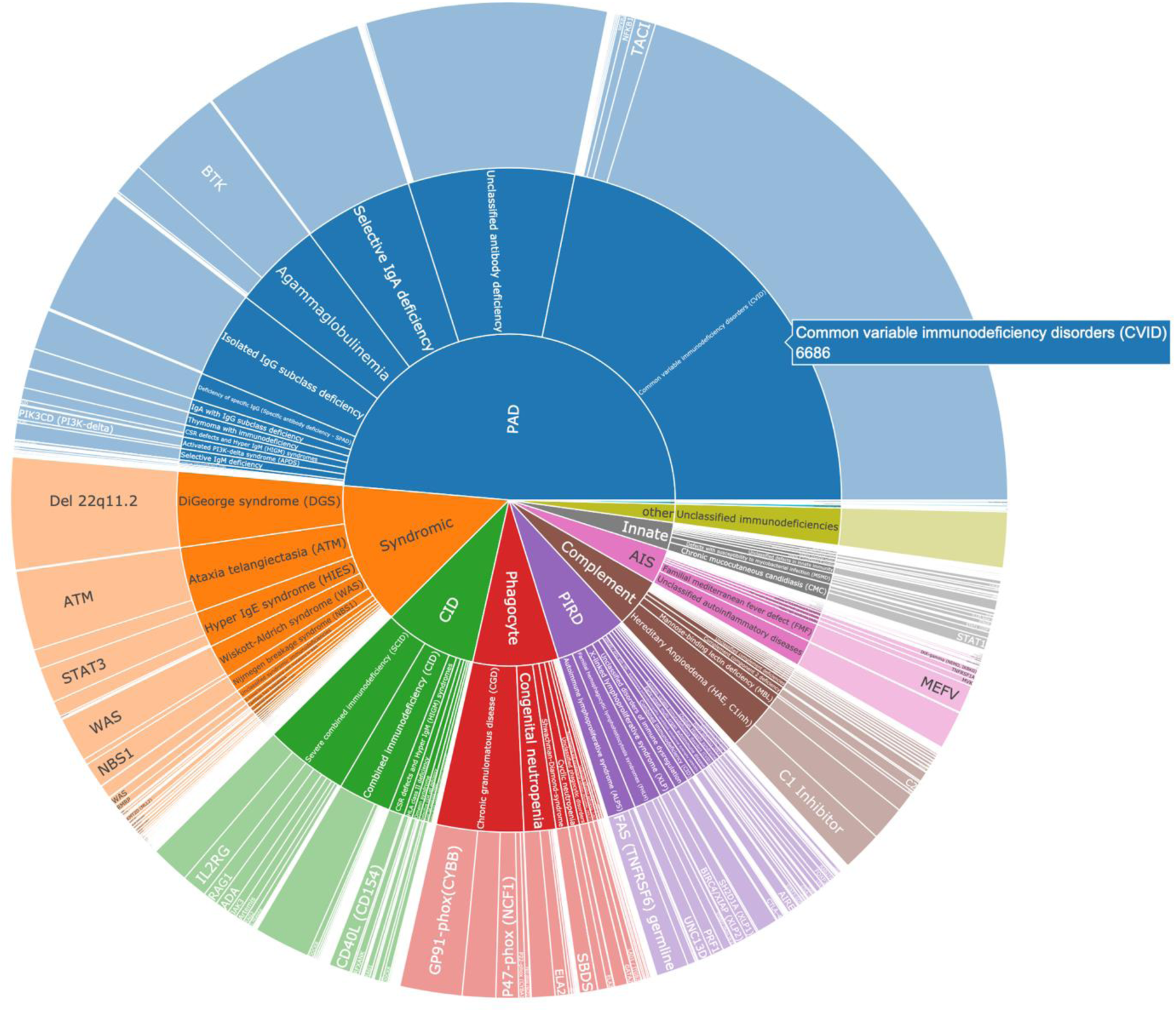
IEI/PID distribution, nested pie chart for IUIS category, diagnosis, and gene. This is merely a sketch, please see the interactive version of the figure with all data labels and patient numbers shown on mouse-over at https://esid.org/html-pages/Suppl%20Fig%203_ESID_30k_sunburst_PID.html.

**Supplementary Figure 4.**
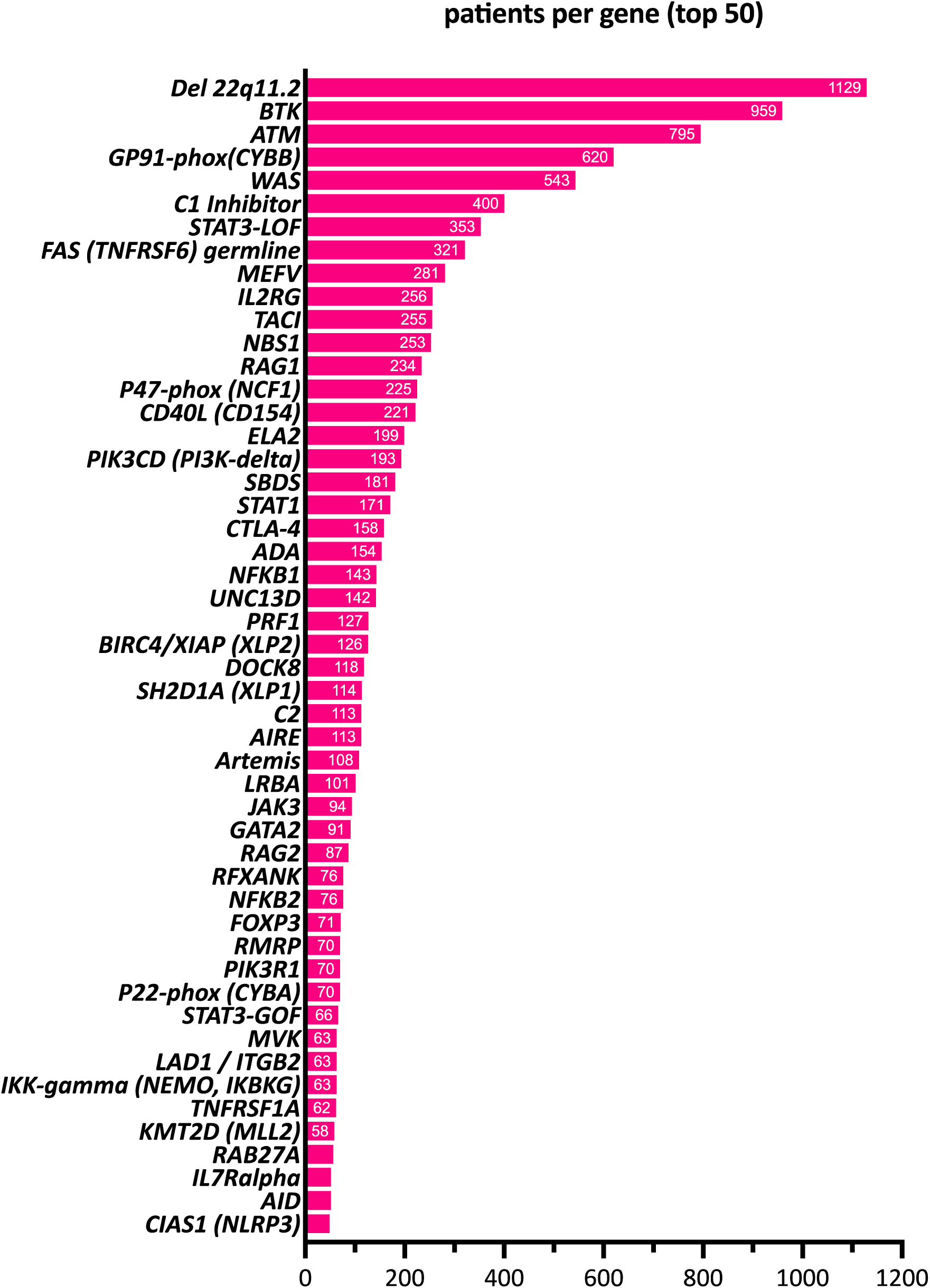
Top 50 genetic diagnoses of IEI/PID patients recorded in the ESID-R. The top 50 genes mutated in ESID-R patients with monogenic IEI/PID are shown with respective patient numbers in descending order.

**Supplementary Figure 5.**
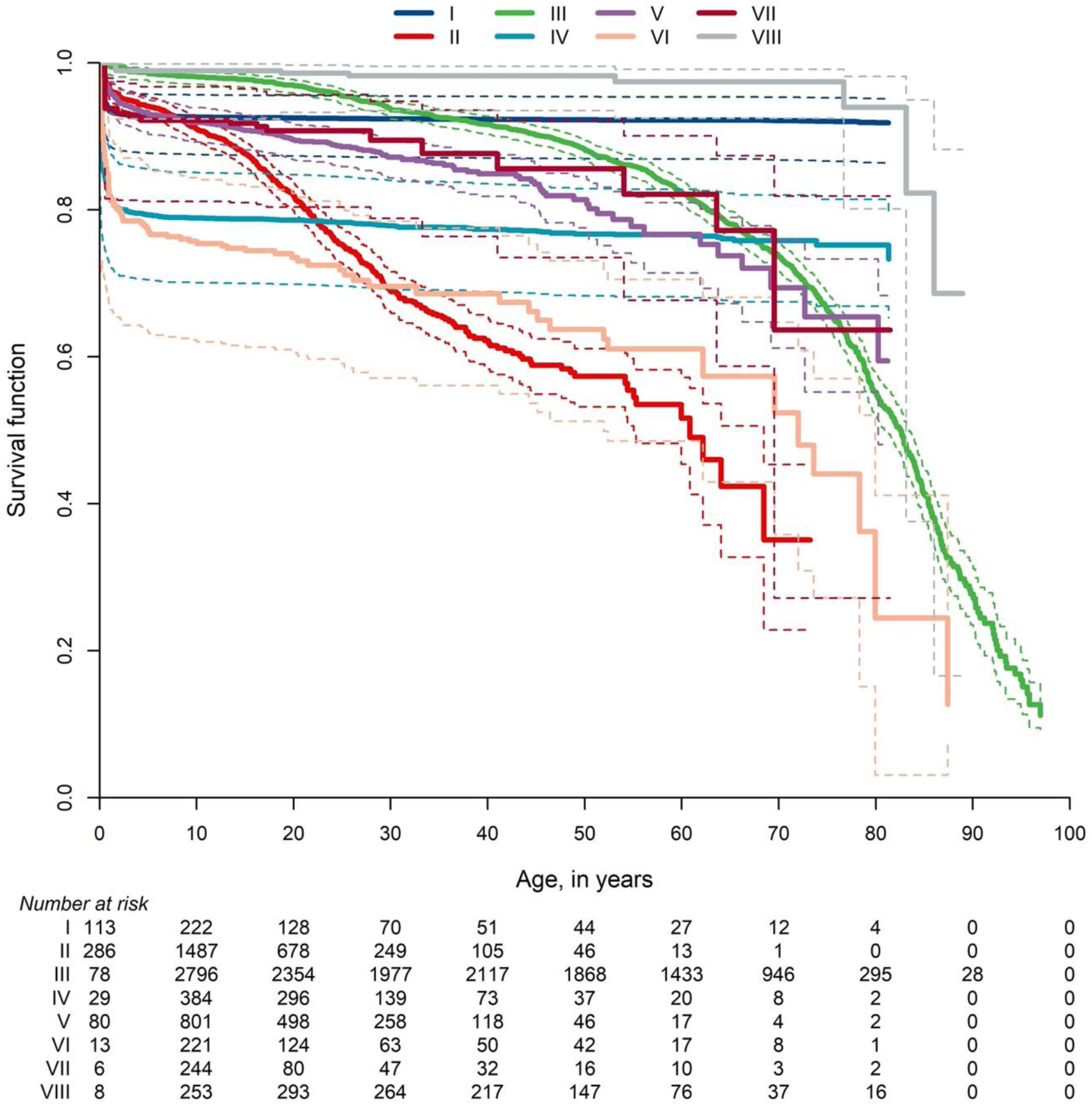
Survival probabilities of main categories of IEI/PID and the living status at last news. Inverse cumulative incidence curves and confidence intervals as described and referenced above. Start = age at diagnosis, stop = age at last news, event = living status (0= censored, 1 = deceased first, 2 = curative therapy first). There were 21,206 patients censored, 1,960 patients who deceased first, and 2,901 patients who had a curative therapy first (not showing deaths after curative therapy); roman numbers refer to the IUIS categories for IEI/PID as listed in Figure 2B.

**Supplementary Figure 6.**
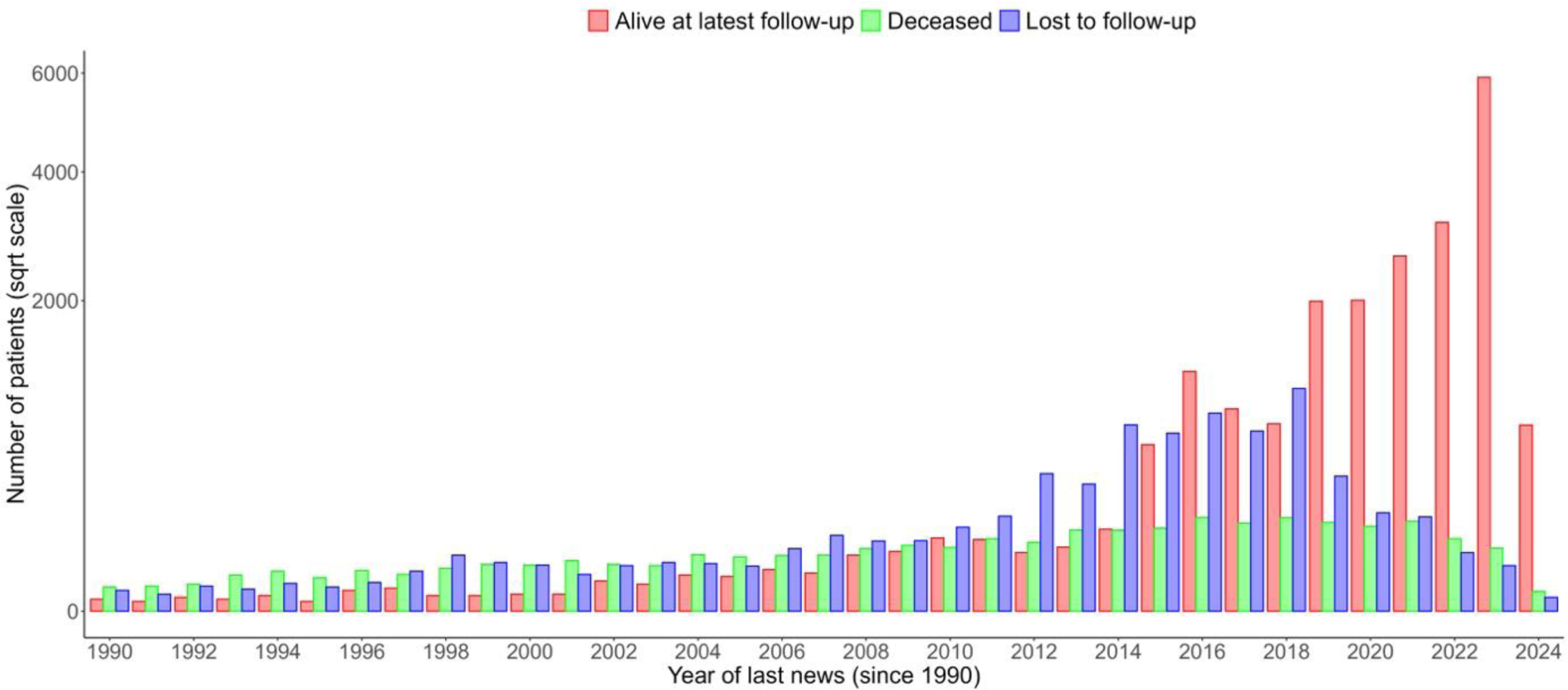
Living status of patients at the year of last news in the ESID-R captured until March 2024. “Last news” is the date when the patient was last seen or his/her condition was reported (by telephone or medical report). It is not the documentation date. Together, these data reflect the overall follow-up rate and living status of patients up to the time point of data closure in March 2024.

**Supplementary Figure 7.**
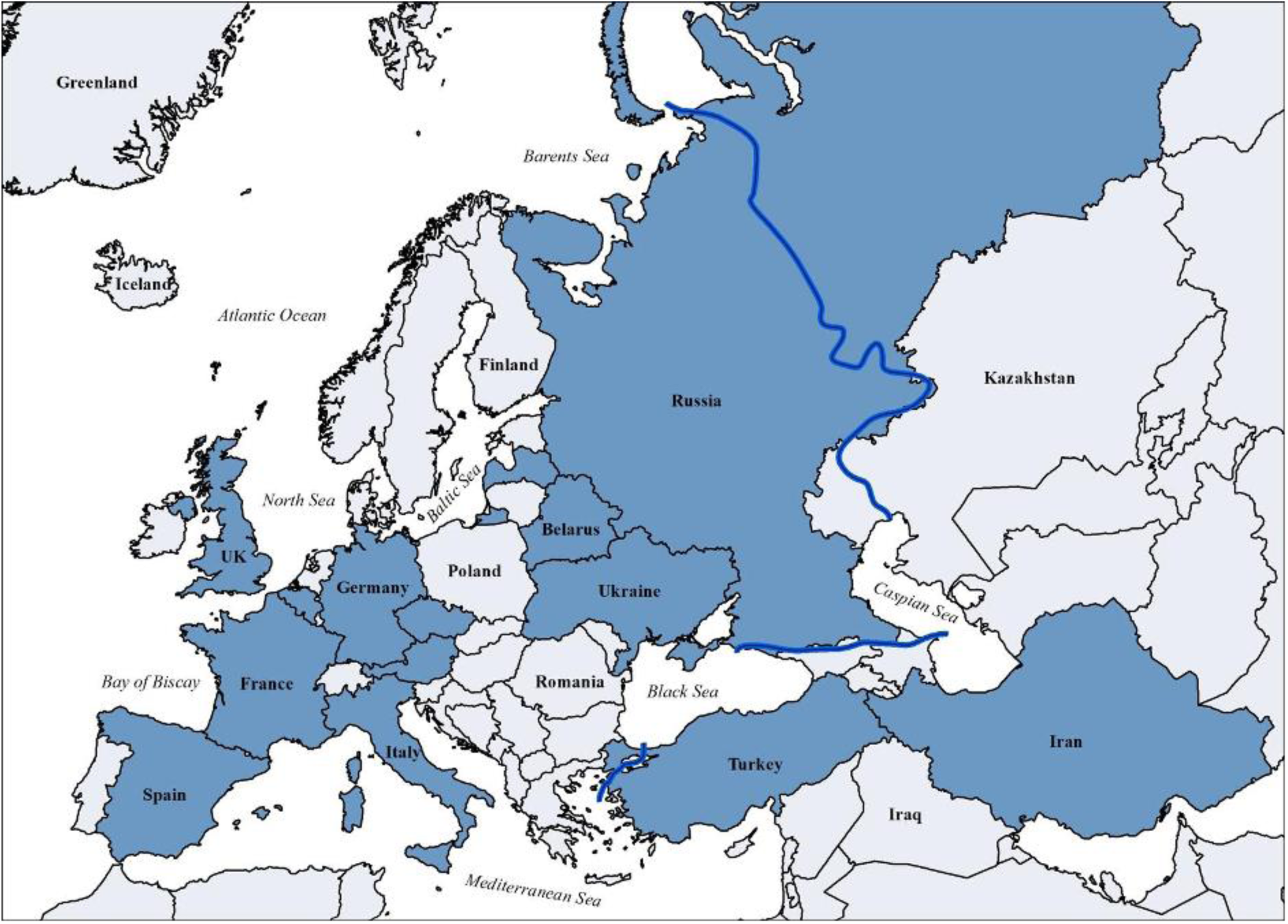
Countries with national patient registries or national sub-registries within the ESID-R according to the 2024 ESID survey (all participating IEI/PID centers worldwide are listed in Suppl. Table 5).

